# Developmental Trajectories of Gut Microbiota and Microbial Metabolites in Children with Autism Spectrum Disorder

**DOI:** 10.64898/2026.05.18.26353458

**Authors:** Federica Del Chierico, Matteo Scanu, Federica Rapisarda, Anna Di Maio, Chiara Marangelo, Pamela Vernocchi, Francesco Errico, Silvia Guerrera, Giovanni Valeri, Alessandro Usiello, Stefano Vicari, Lorenza Putignani

**Affiliations:** Research Unit of Microbiome, Bambino Gesù Children’s Hospital, IRCCS, 00165 Rome, Italy; Department of Environmental, Biological and Pharmaceutical Sciences and Technologies, Università degli Studi della Campania “Luigi Vanvitelli”, 81100 Caserta, Italy; CEINGE Biotecnologie Avanzate “Franco Salvatore”, 80131 Naples, Italy; Department of Agricultural Sciences, University of Naples “Federico II”, 80055 Portici, Italy; Child and Adolescent Neuropsychiatry Unit, Bambino Gesù Children’s Hospital, IRCCS, 00165 Rome, Italy; Life Sciences and Public Health Department, Università Cattolica del Sacro Cuore, 00168 Rome, Italy; Unit of Microbiomics and Unit of Microbiome, Bambino Gesù Children’s Hospital, IRCCS, 00146 Rome, Italy; Department of Life Sciences, Health and Health Professions, Link Campus University, 00165 Rome, Italy

**Keywords:** Autism Spectrum Disorder (ASD), Gut microbiota, Microbial dysbiosis, Neurodevelopment, Short-chain fatty acids (SCFAs), Volatile organic compounds (VOCs), D-amino acids, Microbiome–host interactions, Gut–brain axis, Microbial biomarkers

## Abstract

Autism spectrum disorder (ASD) has been increasingly linked to gut microbial dysbiosis, yet the developmental trajectory of microbiota and microbial metabolites across childhood remains poorly understood. Here, we performed an integrated multi-omics analysis of gut bacterial taxa and faecal metabolites, including volatile organic compounds (VOCs), short-chain fatty acids (SCFAs), and amino acids (AAs), in 101 individuals with ASD and 105 age-matched neurotypical controls. The study was conducted in a well-powered, age-stratified pediatric cohort including toddlers, children, and adolescents.

Our findings reveal early-life microbial dysbiosis in ASD, characterized by elevated alpha-diversity, altered community composition, and age-specific enrichment of taxa including *Faecalibacterium*, *Bacteroides*, *Ruminococcus*, *Alistipes*, and *Roseburia*. Network analyses demonstrated that toddlers exhibit smaller, sparser microbial interaction networks, while children and adolescents show increasingly complex and interconnected networks, compared with respective age-matched controls, suggesting a critical developmental window for microbiome-host interactions.

Metabolomic profiling identified consistent ASD-associated alterations, including elevated aromatic and indole-derived VOCs (i.e., N-ethyl-benzenamine, 6-methyl-5-hepten-2-one, methyl isobutyl ketone), disrupted SCFA patterns, and reduced D-aspartate and D-alanine, indicating functional reprogramming of microbial fermentation and D-amino acid metabolism. Correlations between specific taxa and metabolites suggest that microbial community structure drives these metabolic outputs, with potential impacts on gut-brain signalling, immune modulation, and neurobehavioral phenotypes.

These results support a model in which ASD involves early-life gut microbiota alterations that persist but evolve across development, highlighting potential microbial and metabolic biomarkers. Importantly, they underscore the translational potential of microbiome-targeted interventions during early childhood to modulate neurodevelopmental outcomes.

## 1. Introduction

Autism spectrum disorder (ASD) is a heterogeneous neurodevelopmental condition characterized by social-communication deficits and restricted, repetitive behaviours, with onset in early childhood (Hodges et al., 2020).

Beyond its core behavioural features, ASD is frequently accompanied by gastrointestinal disturbances, immune dysregulation, and metabolic alterations, suggesting the involvement of systemic biological pathways beyond the central nervous system (Puricelli et al., 2022).

In recent years, the gut microbiota has emerged as a key interface between environmental exposures and neurodevelopment, positioning the microbiota–gut–brain axis as a central framework for understanding ASD pathophysiology (Cryan et al., 2019; Sharon et al., 2019).

The human gut microbiota undergoes profound changes during early life, progressively increasing in richness, complexity, and functional capacity from infancy through adolescence. This maturation process is shaped by delivery mode, diet, host genetics, and immune development, and is considered essential for normal metabolic and neuroimmune programming (Bäckhed et al., 2015; La Rosa et al., 2014; Yatsunenko et al., 2012). Disruptions during this critical developmental window may have long-lasting consequences, as early-life microbial perturbations have been shown to influence immune education, intestinal barrier function, and neural signalling pathways (Cryan et al., 2019; Voreades et al., 2014).

Multiple studies have reported alterations in the gut microbiota (i.e., dysbiosis) of individuals with ASD; however, findings regarding microbial diversity and taxonomic shifts remain inconsistent. While some reports describe reduced alpha-diversity or depletion of specific beneficial taxa, others observe increased diversity or no significant differences, likely reflecting variability in age, diet, geography, cohort size, and analytical approaches (Caputi et al., 2024; Chamtouri et al., 2023; Chen et al., 2025; Korteniemi et al., 2024; Scanu et al., 2024; Vernocchi et al., 2022; Yang et al., 2025). Importantly, most studies have focused on taxonomic composition alone, often overlooking the functional outputs of the microbiome that more directly mediate host–microbe interactions (Kadiyska et al., 2025; Novau-Ferré et al., 2025; Toscano De Oliveira et al., 2025; Xiang et al., 2025).

Microbial metabolites, including short-chain fatty acids (SCFAs), volatile organic compounds (VOCs), represent key biochemical mediators of gut–brain communication. SCFAs such as acetate, propionate, and butyrate regulate epithelial integrity, immune responses, and neuroinflammatory processes, and have been implicated in ASD-related behavioral phenotypes (Koh et al., 2016; Parada Venegas et al., 2019). Likewise, microbial fermentation of aromatic amino acids and complex carbohydrates generates VOCs and other small molecules capable of modulating redox balance, mucosal signaling, and neuromodulatory pathways (Louis and Flint, 2017; Parada Venegas et al., 2019; Vernocchi et al., 2022). Although the possible role of D-amino acids (D-AAs) in gut-brain communication remains still unclear, recent observations put forward a notable role of D-aspartate metabolism in regulating NMDA receptor–mediated neurotransmission, immune modulation and ASD phenotypes in human and animal models (Di Maio et al., 2025; Errico et al., 2025; Nuzzo et al., 2020).

Another emerging but understudied dimension of microbiome research in ASD concerns microbial community organization. Network-based approaches provide insight into microbial co-occurrence patterns, ecological stability, and functional interdependence, offering information beyond relative abundance alone. Increasing evidence suggests that microbiome maturation is accompanied by denser and more interconnected microbial networks, reflecting enhanced cooperation and niche specialization during childhood and adolescence.

Given these considerations, a comprehensive, age-resolved investigation integrating microbial taxonomy, metabolomic profiles, and ecological network structure is needed to better understand how gut microbiota alterations unfold across development in ASD. In this study, we characterized the gut microbial ecosystem of a large paediatric ASD cohort compared with age-matched neurotypical controls by integrating 16S rRNA gene sequencing, faecal volatilomics, SCFA quantification, D-AA profiling, Machine Learning (ML) classification, and network analyses. By stratifying subjects into toddlers, children, and adolescents, we aimed to test the hypothesis that ASD is associated with an altered trajectory of microbiota maturation, with early-life dysbiosis representing a critical window of vulnerability that persists, but functionally evolves, across childhood.

## 2. Methods

### 2.1. Study Design, Participant Recruitment, and Sample Collection

Children with ASD and age-/sex-matched neurotypical controls (CTRLs) were recruited at the Bambino Gesù Children’s Hospital (Rome, Italy). The study was approved by the OPBG Ethics Committee (prot. n. 2996_OPBG_2022) and conducted according to Good Clinical Practice and the Declaration of Helsinki. Written informed consent was obtained from all participants.

ASD diagnosis followed the Diagnostic and Statistical Manual of Mental Disorders (DSM-5) criteria and was confirmed using the Autism Diagnostic Observation Schedule (ADOS-2) and the Autism Diagnostic Interview – Revised (ADI-R). CTRLs were selected during a gut microbiota programming survey at the OPBG Microbiome Unit (Vernocchi et al., 2022).

Stool samples were collected at home and stored at −80 °C within 6 hours of collection.

Collected anamnestic and clinical data included demographic and anthropometric measures; neuropsychological characteristics, including autism symptom severity (i.e., low, moderate, or high); behavioural problems, defined as the presence of risk for behavioural difficulties or the absence of clinical symptoms according to the Child Behavior Checklist scales (CBCL-INT, Internalizing Problems; CBCL-EXT, Externalizing Problems; and CBCL-TOT, Total Problems); cognitive level, expressed as IQ/DQ (intelligence quotient/developmental quotient), with or without cognitive impairment or developmental delay; pharmacological treatment; and the presence of epilepsy (Marangelo et al., 2024). The severity of ASD symptomatology was determined by the Calibrated Severity Scores scale (CSS) of the ADOS-2 (Esler et al., 2015; Gotham et al., 2009; Hus and Lord, 2014). Based on the CSS, the patients were divided into two levels of severity according to the ADOS scores criteria (i.e., mild/moderate and severe autism) (American Psychiatric Association, 2013).

### 2.2. DNA Extraction and 16S rRNA Gene Sequencing

Bacterial DNA was isolated from stool samples using a QIAmp Fast DNA Stool Mini Kit (Qiagen, Hilden, Germany), and quantified with a NanoDrop™ 2000/2000c Spectrophotometer (Thermo Scientific, Wilmington, MA, USA).

The V3–V4 regions of the 16S rRNA gene were amplified using the primers 16S_F (5′-TCG TCG GCA GCG TCA GAT GTG TAT AAG AGA CAG CCT ACG GGN GGC WGC AG-3′) and 16S_R (5′-GTC TCG TGG GCT CGG AGA TGT GTA TAA GAG ACA GGA CTA CHV GGG TAT CTA ATC C-3′), as described in the MiSeq rRNA amplicon sequencing protocol (Illumina, San Diego, CA, USA).

PCR was performed at 95 °C for 30 s, 55 °C for 30 s, and 72 °C for 30 s, for 32 cycles, using the Fast Start HiFi Taq kit (Roche Diagnostics, Mannheim, Germany) was used for PCR.

Amplicons were purified with KAPA beads (Roche Diagnostics, Mannheim, Germany), indexed with Nextera primers (Illumina, San Diego, CA, USA), re-purified, quantified using the Quant-iT™ PicoGreen® dsDNA Assay Kit (Thermo Fisher Scientific, Waltham, MA, USA), pooled, and sequenced on the Illumina MiSeq™ platform (Illumina, San Diego, CA, USA).

Raw reads were processed in QIIME2 (Bolyen et al., 2019): paired reads were denoised and merged at 99% similarity using the q2-DADA2 to obtain amplicon sequence variants (ASVs) (Callahan et al., 2016). Taxonomic assignment was performed using q2-Greengenes 2 and the Greengenes database (v2022.10) (v2022.10) (McDonald et al., 2024).

At genus level, abundances were normalized by Cumulative Sum Scaling and transformed to relative values (0–1). For univariate and multivariate analyses, only taxa with relative abundance ≥ 0.01 and present in ≥ 25% of samples in the smallest group were retained to enhance robustness and reduce noise.

### 2.3. Gut Microbiota Metabolomic Profiling by VOCs Analysis

Faecal samples from 97 ASD patients were analysed by GC–MS to detect VOCs by using a carboxen-polydimethylsiloxane coated fiber (85 μm) and a manual solid-phase micro-extraction holder (Supelco Inc., Bellefonte, PA, United States) as previously described (Vernocchi et al., 2020).

### 2.4. Amino Acid Quantification by HPLC

Faecal samples were homogenised (1:20, w/v) in phosphate-buffered saline (PBS), sonicated (3 × 20 s), incubated on ice for 15 min, and centrifuged at 4°C (12,100 g, 5 min) (Gonda et al., 2023). The supernatant was diluted 1:10 in HPLC-grade methanol and centrifuged (13,000 g, 10 min). It was then dried and resuspended in 0.2 M trichloroacetic acid (TCA), while protein pellets were stored at −80 °C for quantification (Serra et al., 2023). The TCA supernatant was neutralised with NaOH and derivatised with o-phthaldialdehyde/N-acetyl-L-cysteine. Amino acid derivatives were separated using a UHPLC Nexera X3 system (Shimadzu) equipped with a Shim-pack GIST C18 column (3 μm, 4.0 × 150 mm) under isocratic conditions (0.1 M sodium acetate buffer, pH 6.2, 1% tetrahydrofuran; flow rate 1 mL/min). Each run was followed by washing (0.1 M sodium acetate, 3% tetrahydrofuran, 47% acetonitrile). Amino acids were identified and quantified by retention times and peak areas relative to internal standards. Total amino acids were normalised to faecal weight (nmol/g faeces) or protein content in brain homogenates (nmol/mg protein). D-Asp/total Asp and D-Ser/total Ser ratios were expressed as percentages, and L-Gln/L-Gln as an absolute value (Di Maio et al., 2025).

### 2.5. Statistical analysis

Alpha- and beta-diversity were calculated on the ASV matrix rarefied to the minimum sequencing depth. Alpha-diversity was compared using the Mann–Whitney U test, and beta-diversity by Permutational Multivariate Analysis of Variance (PERMANOVA).

Differential taxa were identified using the Linear Discriminant Analysis Effect Size (LEfSe) method (Segata et al., 2011) (significance threshold: α = 0.05 and log LDA score > 3.0). Multivariate analyses included Partial Least Squares Discriminant Analysis (PLS–DA) (mixOmics v6.28) and Principal Component Analysis (PCA) (ropls v1.36, Bioconductor).

Using the same normalised genus matrix, three supervised Machine Learning (ML) models—Random Forest (RF), Gradient Boosting Machine (GBM), and Support Vector Machine (SVM)—were applied. Model performance was assessed by the Area Under the Receiver Operating Characteristic (AUROC) curve, reporting sensitivity, specificity, and 95% confidence intervals (CIs) for AUC values.

Correlation analyses were conducted using the Spearman’s rank correlation coefficient. Significant correlations (*p*–value <0.05; |ρ|> 0.6) were visualised as network graphs using the igraph and ggraph (v2.1.1 and v2.2.1, respectively).

Untargeted metabolomic analyses included metabolites detected in ≥10% of samples. Group differences were evaluated by log2 Fold Change (|log2FC| > 1; p < 0.05). PLS–DA and ML models were applied to identify discriminant metabolites. Features consistently selected by ML and significant in univariate analysis (p < 0.05) were retained for downstream analyses. Associations with neuropsychological measures (CSS, CBCL, IQ/DQ) were tested by linear regression and Spearman correlation. Additional correlations were performed between significant metabolites, L/D–AAs, and bacterial genera. SCFAs and L/D–AAs were compared using the Mann–Whitney U test.

Multiple testing was controlled by FDR (Benjamini–Hochberg), unless otherwise specified.

## 3. Results

### 3.1. Cohort Comparison: ASD versus Neurotypical CTRL

A total of 101 ASD and 105 neurotypical CTRL paediatric participants were included, with comparable age distributions (ASD: median 7 years [IQR: 4-9 years]; CTRL: median 8 years [IQR: 4-11 years]). Gender distribution differed (ASD: 79% male; CTRL: 50% male), reflecting the male-to-female prevalence ratio of 3.8 (Maenner et al., 2023). Metataxonomic analysis via 16S rRNA sequencing was performed for all faecal samples. Targeted and untargeted metabolomics were conducted on subsets due to sample availability and quality, ensuring representation across age groups (**Figure S1**).

For 43 ASD participants, demographic, clinical, and cognitive characteristics were obtained (Table 1). Males predominated across all age groups, although the proportion of females increased with age. Autism severity categories (i.e., CSS) showed a higher prevalence of mild–moderate autism in children and a relatively higher proportion of severe autism in toddlers. CBCL-INT, CBCL-EXT, CBC-TOT scores indicated substantial behavioural difficulties, especially in toddlers and adolescents. Cognitive functioning was heterogeneous, with most participants presenting impairment, particularly among children and adolescents, highlighting marked neurodevelopmental variability. No significant differences were observed between groups for CSS total scores, CBCL-INT, CBCL-EXT, CBCL-TOT, or IQ/DQ. Fisher’s exact tests confirmed no differences in categorical clinical variables, indicating comparable symptom severity and cognitive impairment across age groups.

**Table 1.** Demographic, clinical, and cognitive characteristics of the ASD patients stratified for age.

| **Features** |  | **Toddlers** | **Children** | **Adolescents** |
| --- | --- | --- | --- | --- |
|  | Number | 16 | 20 | 7 |
| **Gender** | Male | 13 (81.2%) | 15 (75%) | 4 (57%) |
|  | Female | 3 (19%) | 5 (25%) | 3 (43%) |
| **Age** | mean ± SD | 5 ± 1 | 7 ± 1 | 12 ± 3 |
| **CSS total score** | mean ± SD | 7 ± 2 | 7 ± 1 | 8 ± 2 |
| **CSS categories** | mild-moderate autism | 6 (37%) | 15 (75%) | 4 (57%) |
|  | severe autism | 7 (44%) | 5 (25%) | 2 (29%) |
|  | not available | 3 (19%) | 0 (0%) | 1 (14%) |
| **CBCL internalizing score** | mean ± SD | 66 ± 7 | 63 ± 9 | 68 ± 11 |
| **CBCL internalizing** | no symptoms | 2 (13%) | 5 (25%) | 1 (14%) |
|  | symptoms | 6 (37%) | 9 (45%) | 4 (58%) |
|  | at risk | 6 (37%) | 3 (15%) | 1 (14%) |
|  | not available | 2 (13%) | 3 (15%) | 1 (14%) |
| **CBCL externalizing score** | mean ± SD | 59 ± 9 | 55 ± 9 | 60 ± 5 |
| **CBCL externalizing** | no symptoms | 5 (31%) | 11 (55%) | 2 (29%) |
|  | symptoms | 6 (37%) | 3 (15%) | 1 (14%) |
|  | at risk | 3 (19%) | 3 (15%) | 3 (43%) |
|  | not available | 2 (13%) | 3 (15%) | 1 (14%) |
| **CBCL total problems score** | mean ± SD | 67 ± 8 | 60 ± 9 | 68 ± 7 |
| **CBCL total problems** | no symptoms | 4 (25%) | 9 (45%) | 1 (14%) |
|  | symptoms | 10 (63%) | 6 (30%) | 4 (58%) |
|  | at risk | 0 (0.0%) | 2 (10%) | 1 (14%) |
|  | not available | 2 (12%) | 3 (15%) | 1 (14%) |
| **IQ/DQ score** | mean ± SD | 65 ± 27 | 60 ± 11 | 60 ± 22 |
| **IQ/DQ** | with cognitive impairment | 9 (56%) | 15 (75%) | 5 (71%) |
|  | without cognitive impairment | 6 (38%) | 5 (25%) | 2 (29%) |
|  | not available | 1 (6%) | 0 (0%) | 0 (0%) |

### 3.2. Gut Microbiota Composition and Diversity

Gut microbiota analysis showed significantly higher alpha-diversity (Shannon–Wiener, Simpson) in ASD versus CTRL, indicating a richer, more heterogeneous community (**Figure S2A and B**).

Beta-diversity analyses revealed significant compositional differences between ASD and CTRL microbiota profiles (PERMANOVA p-values: Bray–Curtis, p = 0.001; Unweighted UniFrac, p = 0.001) (**Figure S2C and D**), demonstrating robust separation between diagnostic groups.

Several taxa showed significantly higher relative abundance in ASD, including *Phocaeicola*_A_858004, *Faecalibacterium*, *Bacteroides*_H, *Alistipes*_A_871400, *Prevotella*, *Dialister*, *Parabacteroides*_B_862066, *Roseburia*, *Dysomobacter*, *Ruthenibacterium*, *Eisenbergiella*, and *Barnesiella* (**Figure S2E**).

Conversely, ASD subjects showed reduced abundances of *Bifidobacterium*_338775, *Akkermansia*, *Blautia*_A_141781, *Streptococcus*, *Collinsella*, *Anaerostipes*, and *Agathobacter*_164117 (**Figure S2E**).

Three ML models (RF, GBM, SVM) discriminated ASD from CTRL at genus level (AUCs: RF 0.85, GBM 0.82, SVM 0.69), indicating that microbial taxonomic profiles provide moderate-to-strong discriminative power (**Figure S2F and Figure S3**).

Combining the ML and univariate results, we filtered out *Ruminococcus*_B and *Eubacterium*_R as they were not statistically significant in the univariate analysis. The remaining genera represent a consistent microbial signature, distinguishing ASD from CTRL: *Faecalibacterium*, *Bacteroides*_H, *Dialister*, *Phocaeicola*_A_858004, *Streptococcus*, *Alistipes*_A_871400, *Roseburia*, *Ruthenibacterium*, *Prevotella*, *Bifidobacterium*_338775, and *Blautia*_A_141781.

Overall, ASD patients show altered microbial diversity and structure, with higher alpha-diversity, distinct beta-diversity, multiple differentially abundant genera, and strong ML-based discriminative power, supporting gut microbiota as potential ASD biomarkers.

### 3.3. Age-stratified Analyses

To assess age-dependent microbial signatures in ASD, participants were stratified as toddlers (1–5 y: 44 ASD, 36 CTRL), children (6–10 years: 38 ASD, 32 CTRL), and adolescents (>10 years: 19 ASD, 37 CTRL). Comparisons were performed within ASD and CTRL groups and between groups by age class.

Alpha- and beta-diversity analyses showed no significant differences across ASD age groups (**Figure S4A and B, S4E and F**), suggesting relative microbiota stability.

The same result was reported for alpha-diversity analysis in CTRL age groups (**Figure S4C and D**), while the beta-diversity analysis showed significant compositional differences amongst CTRL age group (PERMANOVA p-values: Bray–Curtis, p = 0.001; Unweighted UniFrac, p = 0.001) (**Figure S4G and H**). This result demonstrated a normal developmental maturation of the gut microbiota in the CTRL population, while ASD exhibited a stronger disease-associated signature. ASD toddlers were enriched in *Faecalibacterium*, *Ruminococcus*_E, and *Eisenbergiella*, children in Alistipes_A_871404, *Hungatella*_A_128155, *Ruminococcus*_C_58660, *Barnesiella*, and CAG-217, while adolescents in ER4 (Figure S2A) (**Figure S4I)**.

CTRLs showed age-specific increases in several taxa: *Bifidobacterium*_388775, *Streptococcus*, *Blautia*_A_141781, *Erysipelatoclostridium*, *Ruminococcus*_B, *Eggerthella*, *Anaerostipes*, and *Mediterraneibacter*_A_155507 in toddlers; *Collinsella*, *Dysosmobacter*, *Ruthenibacterium*, *Anaerobutyricum*, *Copromorpha*, *Dorea*_A, *Faecousia*, and ER4 in children; *Akkermansia*, *Bacteroides*_H, *Gemmiger*_A_73129, *Phocaeicola*_A_858004, *Alistipes*_A_871400, *Flemicutes*, *Agathobacter*_164117, *Barnesiella*, *Eubacterium*_R, CAG-273, *Parabacteroides*_B_862066, CAG-177, *Prevotella,* and *Ruminococcus*_D in adolescents (**Figure S4J)**.

### 3.4. Age-Specific Microbial Taxa Abundance Patterns

Shannon diversity was higher in ASD toddlers and adolescents (**Figure S5A and C**), while Simpson diversity differed only in adolescents (**Figure S5F**). Bray–Curtis and Unweighted UniFrac consistently separated ASD and CTRLs across all ages (**Figure S5G–L**).

ASD toddlers showed increased *Bacteroides*_H, *Faecalibacterium*, *Phocaeicola*_A_858004, *Alistipes*_A_871400, *Gemmiger*_A_73129, *Dialister*, *Ruminococcus*_D, *Prevotella*, *Parabacteroides*_*B*_862066, *Parasutterella*, *Dysomobacter*, *Eisenbergiella*, *Enterocloser*, *Ruthenibacterium*, *Barnesiella*, *Acetatifactor*, *Mediterraneibacter* A_155507, *Agathobacter*_164117, and *Roseburia* and decreased *Bifidobacterium*_338775, *Escherichia*_710834, *Streptococcus*, *Blautia*_A_141781, *Erysipelatoclostridium*, *Eggerthella*, *Anaerostipes*, and *Ruminococcus*_B (**Figure S6A)**. In children *Phocaeicola*_A_858004 and *Parasutterella* increased (**Figure S6B)**; in adolescents *Ruminococcus*_E and *Akkermansia* decreased (**Figure S6C)**.

### 3.5. ML Classification Across Age Groups

In toddlers, all ML models showed strong discriminatory power (AUC > 0.8), with RF and GBM performing best (**Figure 1A and D**); top taxa included *Faecalibacterium*, *Streptococcus*, *Eggerthella*, *Bifidobacterium*_388775, and *Bacteroides*_H. Children showed moderate discrimination (AUC RF = 0.65, GBM = 0.71, SVM = 0.73) (**Figure 1B and E**). In this group, the important taxa were *Faecalibacillus*, *Streptococcus*, *Phocaeicola*_A_858004, *Bacteroides*_H, *Parasutterella*, *Barnesiella*, *Mediterraneibacter*_A_15550, *Escherichia*_710834, *Eubacterium*_R, and *Prevotella*. In adolescents, RF retained fair classification performance (AUC = 0.73) whit key taxa *Faecalibacterium*, *Roseburia*, CAG-41, *Akkermansia*, *Barnesiella*, *Ruminococcus*_E, *Prevotella*, *Alistipes*_A_871400, *Collinsella,* and *Dialister* (**Figure 1C and F**).

**Figure 1.**
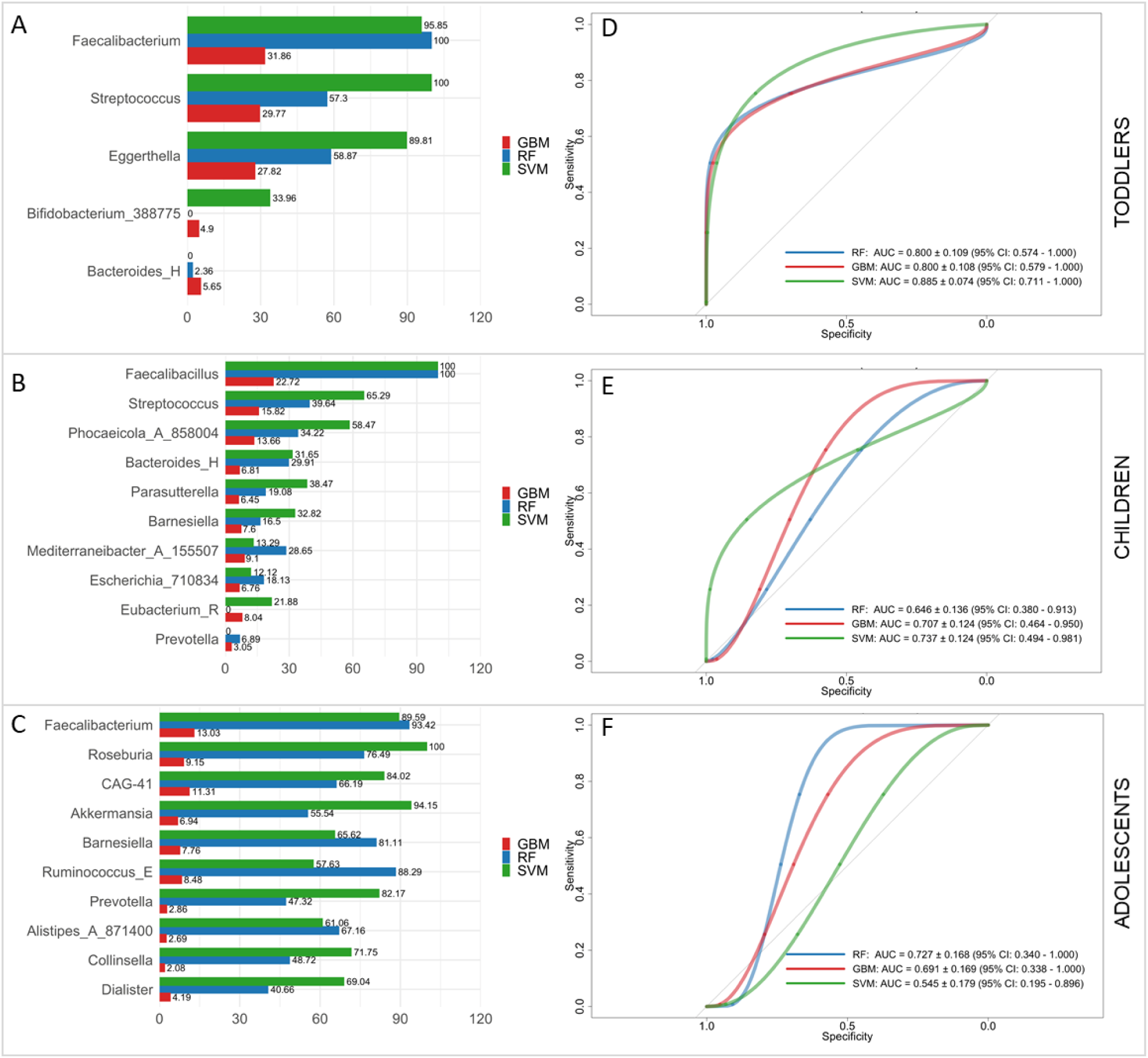
ROC-based Machine Learning (ML) classification performance within each age group for Random Forest (RF), Gradient Boosting Machine (GBM), and Support Vector Machine (SVM). Top predictors are shown for each group: (A) toddlers, AUC ≥ 0.80; (B) children, RF = 0.65, GBM = 0.71, SVM = 0.73; and (C) adolescents, RF = 0.73, GBM = 0.69, SVM = 0.55.

Overall, microbial signatures discriminating ASD from CTRL are strongest in early childhood, becoming less pronounced with age.

### 3.6. Microbial Network Analysis

Network analyses performed using stringent correlation thresholds (|ρ| ≥ 0.6) revealed distinct age-dependent microbial interaction networks in ASD subjects (**Figure 2**). Network topology differed markedly across developmental stages, with progressively greater complexity and connectivity observed from toddlers to children and adolescents.

**Figure 2.**
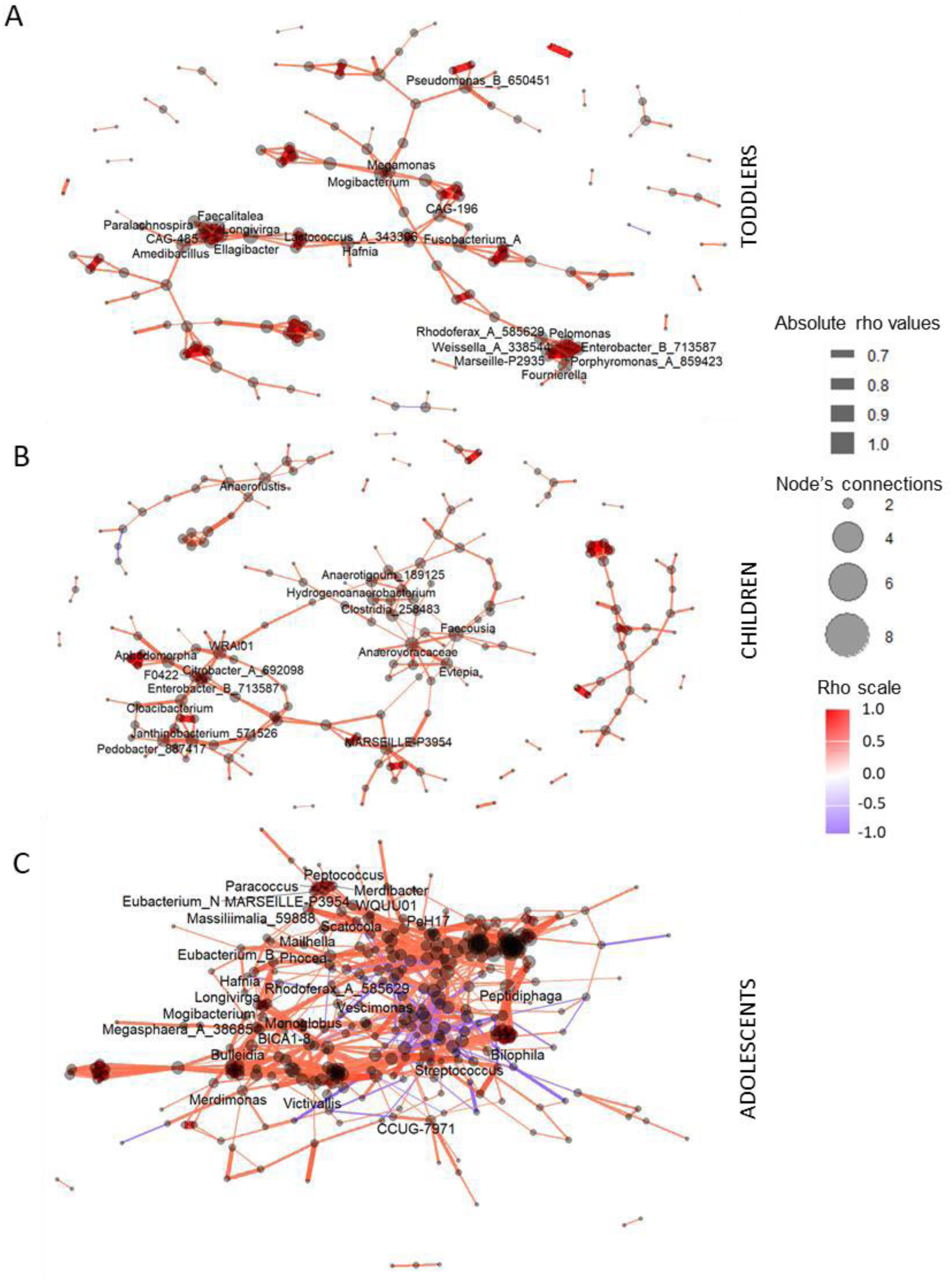
Age-dependent microbial co-occurrence networks in ASD subjects. Microbial interaction networks were constructed separately for toddlers **(A)**, children **(B)**, and adolescents **(C)** with ASD using Spearman correlation analysis with stringent thresholds (|rho| ≥ 0.6). Nodes represent bacterial taxa, with node size proportional to degree (number of connections), while edges represent significant correlations between taxa, coloured according to correlation strength and direction (red, positive; purple, negative). Only taxa with nodes having more than 6 connections are shown.

In ASD toddlers, the network was sparse and weakly connected, comprising 147 nodes and 420 edges, with a low average number of neighbours per node (mean ± SD = 3.86 ± 1.93). In ASD children, the network expanded moderately, with an increased number of nodes (204) but a similar level of local connectivity (mean ± SD = 3.67 ± 1.88), indicating partial consolidation of microbial interactions. In contrast, ASD adolescents exhibited a markedly denser and more complex network, with 309 nodes and 2,616 edges and a substantially higher mean number of neighbours per node (mean ± SD = 9.47 ± 7.55), reflecting extensive microbial interconnectivity.

### 3.7. Untargeted Volatilome Profiling

Volatilome analysis identified 13 metabolites differing between ASD and CTRL (**Figure S7**). Several compounds, including methyl salicylate, 6-Hepten-1-ol, 2-methyl, propanoic acid, ethyl ester, pentanoic acid, ethyl ester, butanoic acid, ethyl ester, 5-Hepten-2-one, 6-methyl-, methyl isobutyl ketone, benzenamine N-ethyl, and benzothiazole were markedly increased in ASD compared with CTRLs. In contrast, metabolites such as methional, 6-Methyl-5-hepten-2-one, 2-Butanone, and 1-Heptanol were significantly higher in neurotypical subjects. No differences were observed within ASD stratified by delivery modality, neonatal feeding, and food selectivity.

### 3.8. Multivariate and ML Analyses

PLS-DA demonstrated a clear separation between ASD and neurotypical metabolic profiles (R² = 0.531; Q² = 0.279; AUROC = 0.886) (**Figure S8A-D**). Metabolites with VIP > 1.2 represented the strongest contributors to group discrimination and substantially overlapped with significant univariate results (**Figure S7**).

ML classifiers on metabolomics achieved high accuracy (RF/GBM 0.96, SVM 0.89) (**Figure 3A-C**). Feature importance rankings revealed substantial concordance across all models. The most consistently discriminant metabolites across algorithms were benzenamine, N-ethyl-, 5-Hepten-2-one, 6-methyl-, and methyl isobutyl ketone, all significantly elevated in ASD. ROC curve analysis for each yielded high AUC values, supporting their potential as candidate biomarkers.

**Figure 3.**
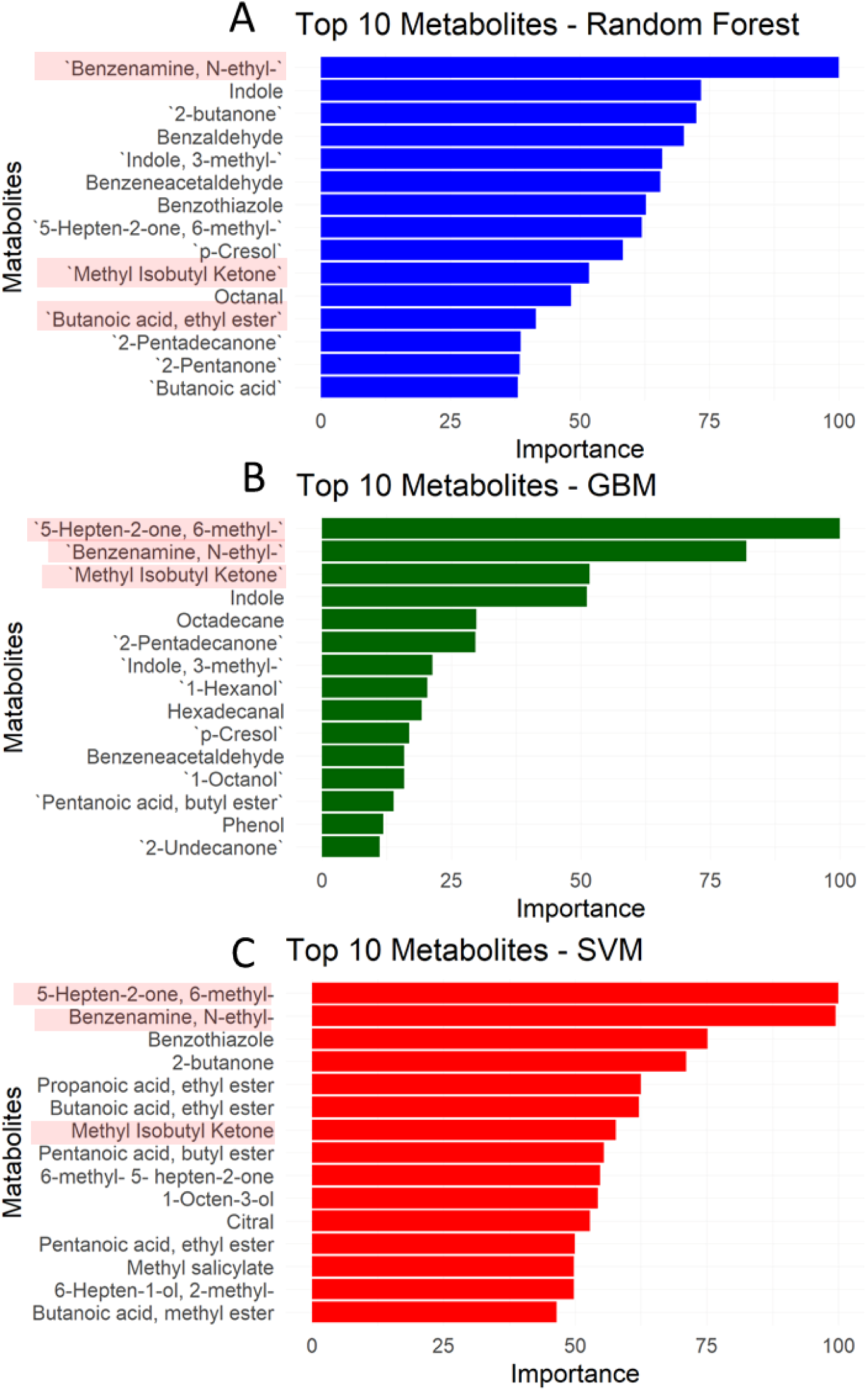
Machine Learning (ML) classification performance on VOC metabolomic profiles in ASD and CTRL. ML models trained on VOC metabolomic profiles exhibit high discriminative power in distinguishing ASD from CTRL subjects. Random Forest (RF) and Gradient Boosting Machine (GBM) models achieve accuracy values of 0.964, recall of 1.00, F1-scores of 0.976, and AUC values between 0.94 and 0.99. Support Vector Machine (SVM) performs slightly lower (accuracy = 0.893, AUC = 0.944) but remains above chance. These results indicate that the VOC metabolic signature is robust, reproducible, and model-independent. Metabolites identified by all three models were highlighted in red in the figure.

The linear regression analysis was applied to the ML-identified metabolites and the ASD behavioural scales. A significant association emerged between CBCL-EXT at-risk scores and benzenamine, N-ethyl- and between CBCL-INT at-risk scores and methyl isobutyl ketone, indicating a diagnostic severity-predictive value of these two molecules (**Figure S9**).

Concentrations of ML-identified metabolites are not associated with cognitive quotient measures and severity scores, indicating that metabolic biomarkers reflect diagnostic classification rather than cognitive impairment (**Figure S10**).

The correlation analysis amongst behavioural scales and total VOCs revealed a significant positive correlation between CSS total score and pentanoic acid ethyl ester, as well as between CBCL-EXT and CBCL-TOT with methyl salicylates (**Figure S11A**). Then, we performed a correlation analysis between these two selected metabolites and bacterial taxa (**Figure S11B**).

Methyl salicylate showed positive associations with CAG-95, *Mediterraneanbacter*_A_155507 and *Moraxella*_C_651924, while displaying negative associations with *Pseudomonas*_E_647464, RUG115, CAG-83, UBA644, *Akkermansia*, *Anaerotignum*_189125, SFA01, and CAG-1427. In contrast, pentanoic acid, ethyl ester exhibited an almost complementary pattern, showing negative associations with *Solibaculum*, *Clostridium*_Q_134516, *Anaerotruncus*, and UBA1417 and a strong positive association with *Haemophilus*_D_35815, *Neisseria*_56320, *Fructilactobacillus*, *Butyricicoccus*_A_77419, *Alloiococcus*, *Scardovia*, *Agathobacter*_164117, *Megasphaera*_A_36885, *Nanosynbacter*, *Oribacterium*, CAULXD01, *Haemophilus*_A, *Eubacterium*_B, *Granulicatella*, and CAG-267.

### 3.9. Age-Specific VOCs Abundance Patterns

ASD toddlers exhibited elevated benzaldehyde, indole, benzeneacetaldehyde, 5-Hepten-2-one, 6-methyl-, P-Cresol, 3-methyl-indole, benzenamine N-ethyl-, and benzothiazole versus CTRL. In children, ethanol levels were increased in CTRL, while adolescents showed no significant differences (**Figure S12)**.

The levels of 5-hepten-2-one, benzenamine N-ethyl and benzothiazole were also higher in the overall ASD group. This means that these compounds are associated with ASD, independently of age.

### 3.10. SCFA Concentrations

Quantitative analysis of SCFAs revealed group-level differences, even if not statistically significant. Across the cohort, individuals with ASD exhibited higher concentrations of acetic and propanoic acids, alongside reduced levels of butanoic and pentanoic acids compared with neurotypical controls (**Table 2**).

**Table 2.** SCFA mean concentrations (mg/kg) in ASD vs CTRL overall groups and by age class groups. P values were calculated by Mann–Whitney test.

| SCFA | OVERALL COHORT |  | P<br>values | TODDLERS |  | P<br>values | CHILDREN |  | P<br>values | ADOLESCENTS |  | P<br>values |
| --- | --- | --- | --- | --- | --- | --- | --- | --- | --- | --- | --- | --- |
|  | CTRL | ASD |  | CTRL | ASD |  | CTRL | ASD |  | CTRL | ASD |  |
| Acetic acid (mg/kg)<br>(Mean ± SD) | 9.07 ±<br>48.58 | 103.14 ±<br>359.5 | 0.28 | 4.93 ±<br>14.80 | 90.28 ±<br>283.54 | 0.73 | 0 | 39.92 ±<br>157.94 | 0.39 | 20.3 ±<br>76.17 | 251.37 ±<br>652.75 | 0.34 |
| Butanoic acid (mg/kg)<br>(Mean ± SD) | 754.03 ±<br>1249.91 | 651.8 ±<br>1286.05 | 0.72 | 157.88 ±<br>233.35 | 532.3 ±<br>940.35 | 0.83 | 667.33 ±<br>1088.18 | 480.75 ±<br>823.65 | 0.51 | 1246.78 ±<br>1619.02 | 1240.07 ±<br>2242.88 | 0.73 |
| Propanoic acid (mg/kg)<br>(Mean ± SD) | 66.14 ±<br>208.67 | 88.69 ±<br>332.81 | 0.35 | 28.17 ±<br>40.79 | 58.22 ±<br>240.51 | **0.025** | 78.58 ±<br>203.46 | 49.64 ±<br>98.2 | 0.61 | 80.95 ±<br>285.85 | 230.05 ±<br>643.37 | 0.36 |
| Pentanoic acid (mg/kg)<br>(Mean ± SD) | 158.72 ±<br>395.58 | 154.87 ±<br>391.61 | 0.56 | 19.61 ±<br>23.94 | 170.5 ±<br>433.2 | 0.94 | 120.06 ±<br>315.67 | 107.24 ±<br>252.47 | 0.61 | 290.71 ±<br>545.44 | 210.55 ±<br>508.07 | 0.79 |

Stratification by developmental stage revealed higher concentrations of all four SCFAs in ASD toddlers relative to age-matched controls, but only propanoic acid was statistically significant (**Table 2**). Children showed elevated acetic acid but decreased butanoic, propanoic, and pentanoic acids, suggesting a shift in the balance of fermentative processes during mid-childhood. Adolescents presented with higher acetic and propanoic acids and reduced butanoic and pentanoic acids, mirroring the global cohort trend (**Figure S13**).

Correlation analysis between bacterial taxa and SCFAs in toddlers showed that butyrate was positively correlated with *Blautia*_A_141781 and *Agathobacter*_164117, and negatively with *Bacteroides*_H. In children, *Parabacteroides_B_862066* was negatively correlated with acetate; *Faecalibacterium* positively correlated with pentanoic acid and butanoic acid and *Phocaeicola_A_858004* with pentanoic acid and acetic acid (**Figure S14**). In adolescents, *Faecalibacterium* was positively correlated with pentanoate and *Dialister, Phocaeicola_A_858004,* and *Bacteroides_H* showed a negative association with acetate (**Figure S14**). These age-dependent patterns suggest a dynamic maturation of microbial-metabolic interactions across childhood in ASD.

### 3.11. Differential Abundance of Faecal AAs

Correlation analyses revealed complex associations between L-/D-AAs and bacterial taxa (**Figure S15**). Overall, the distribution of correlation values spanned a broad range, suggesting that specific bacterial taxa may differentially contribute to, or be influenced by, D-AA concentrations. These findings point to the coexistence of taxa potentially involved in D-AA production and others associated with D-AA depletion or utilization. In particular, *Alistipes*_A_871400, ER4, *Ruthenibacterium*, *Gemmiger*_A_73129, UMGS1071, *Lawsonibacter*, *Terrisporobacter*, *Intestinibacter*, *Hungatella*_A_128155, *Streptococcus*, *Bifidobacterium*_388775, *Fimenecus* and *Eubacterium*_G were correlated with the levels of D-aspartate, D-alanine and D-serine.

In the overall ASD and CTRL groups, quantitative analysis revealed that D-aspartate (**Figure 4A**) and D-alanine (**Figure 4C**) were decreased in the ASD group compared to the CTRL one. Furthermore, this trend was also observed for D-alanine in ASD toddlers (**Figure 4G**) and for D-aspartate in ASD children (**Figure 4F**), compared to their respective CTRL groups.

**Figure 4.**
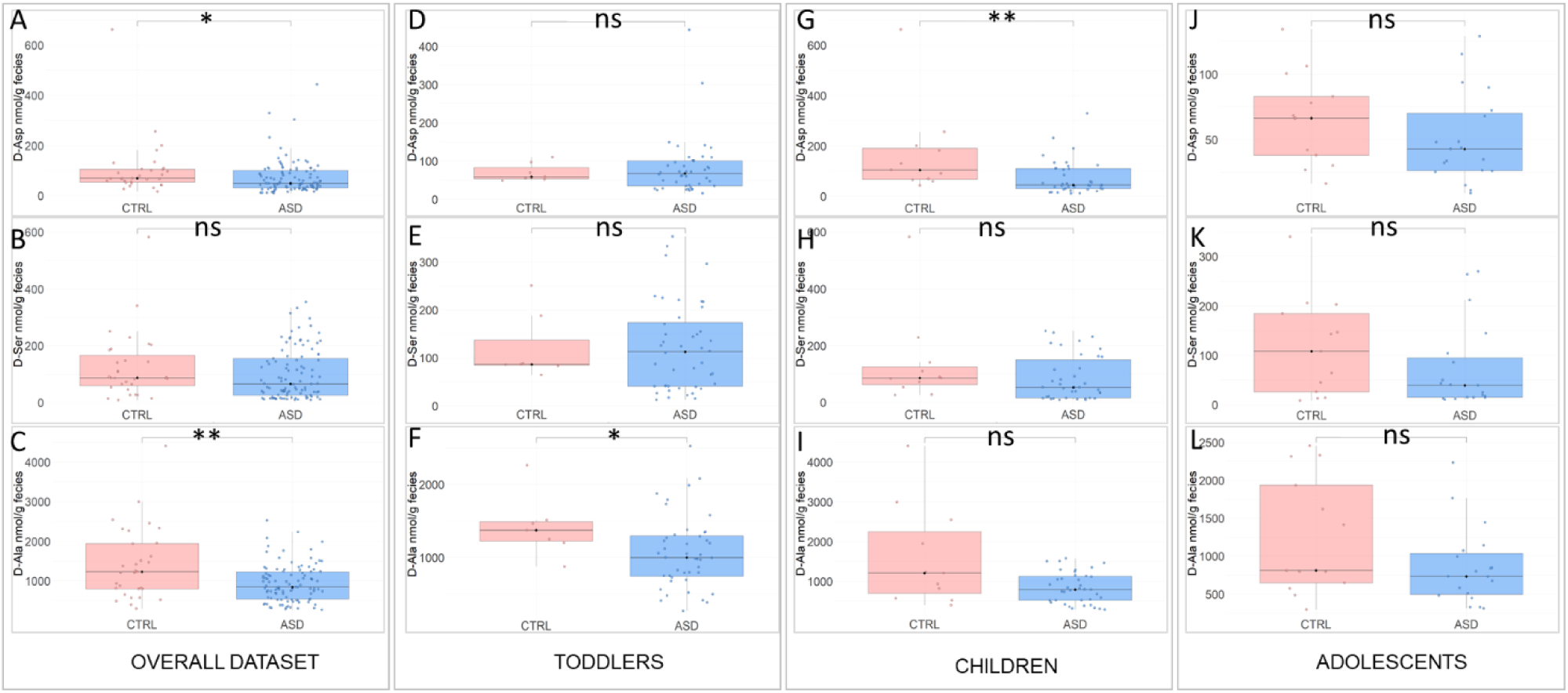
Box plot of D-AA differentiating ASD vs CTRL and within each age group. The box plot shows the distribution of D-Aspartate, D-Serine and D-Alanine in stool samples. * comparisons statistically significant (p_adj_ < 0.05).

Distinct correlation profiles emerged across ages. ASD toddlers showed numerous positive and negative correlations, indicating transitional microbial–AA interactions. Children and adolescents had fewer, mostly positive or negative associations (**Figure S16**). GABA correlated positively with butanoic/propanoic derivatives and negatively with D-serine, D-alanine, and D-aspartate across ages (**Figure S17**), showing stable yet age-specific microbiota-metabolite dynamics.

## 4. Discussion

In this study, we characterized the gut microbial ecology of a large paediatric ASD cohort compared with age-matched neurotypical CTRLs by integrating bacterial taxa and faecal metabolites, including VOCs, SCFAs and AAs, with a specific focus on D-AAs. Our findings provide robust evidence of ASD-associated shifts in microbial structure, function and metabolism, further supporting growing literature on gut dysbiosis in ASD.

A first notable finding was the higher alpha-diversity observed in ASD subjects. While previous studies report variable trends, some describing reduced diversity, others finding no differences, our data indicate a richer and more heterogeneous microbial community in ASD. In our well-powered cohort, increased diversity may reflect ecological instability or compensatory overgrowth of specific bacterial groups in ASD (Marangelo et al., 2024; Scanu et al., 2024; Vernocchi et al., 2022). Nonetheless, factors like age, geography, diet, sample size, and sequencing methodology may have further influenced variability across studies.

Beta-diversity showed clear separation between ASD and CTRLs, suggesting community-wide restructuring. ML models showed strong performance, with overlapping taxa identified by ML and univariate analyses, defining a core ASD-associated signature including *Faecalibacterium*, *Bacteroides*, *Dialister*, *Phocaeicola, Streptococcus, Alistipes*, *Roseburia*, *Ruminococcus*, *Ruthenibacterium*, *Eubacterium*, *Prevotella*, *Bifidobacterium*, and *Blautia*.

Several of these genera are involved in SCFA production or complex carbohydrate fermentation, pathways previously implicated in gut–brain communication. In particular, *Faecalibacterium*, *Roseburia*, *Eubacterium*, *Ruminococcus*, and *Blautia* are recognized as major butyrate-producing taxa (Louis and Flint, 2017; Parada Venegas et al., 2019; Silva et al., 2020). At the same time, *Bacteroides*, *Phocaeicola*, *Prevotella*, *Bifidobacterium*, and *Alistipes* play key roles in the degradation of dietary fibers and polysaccharides, providing substrates for SCFA synthesis through microbial cross-feeding (Flint et al., 2012; Koh et al., 2016; Turroni et al., 2018). *Dialister* and *Streptococcus* contribute indirectly through lactate and intermediate metabolite production, whereas the functional role of *Ruthenibacterium* remains less well characterized (Belenguer et al., 2006; Louis and Flint, 2017). SCFAs regulate barrier integrity, immune and neuroinflammatory responses, and neural signalling, implicating the microbiota–gut–brain axis (Cryan et al., 2019; Dalile et al., 2019), and consistent alterations in SCFA levels were also observed in our metabolomic analyses.

Enrichment of *Prevotella* and *Dialister*, linked to dietary patterns and mucosal immune activation, suggests possible alterations in metabolic and inflammatory microenvironments in ASD (Iljazovic et al., 2021; Socała et al., 2021; Valencia-Buitrago et al., 2025).

Age-stratified analyses revealed further insights. Whereas neurotypical microbiota undergo marked age-dependent maturation with increasing alpha-diversity and community differentiation with age, ASD microbiota remained comparatively stable, with alterations detectable across developmental stages (Bäckhed et al., 2015; La Rosa et al., 2014; Stewart et al., 2018; Yatsunenko et al., 2012). This pattern suggests a potentially altered maturation trajectory, possibly linked to dietary rigidity, gastrointestinal symptoms, or neurobehavioral factors. Within ASD, taxonomic composition varied by age. Toddlers with ASD showed increases in *Faecalibacterium*, *Ruminococcus*_E, and *Eisenbergiella*, whereas ASD children were characterized by enrichment of taxa such as *Alistipes*, *Hungatella*, *Ruminococcus*, *Barnesiella,* and CAG-217. In ASD adolescents, ER4 was the most distinctive taxon. Thus, microbial composition evolves with development, yet ASD–CTRL differences persist, indicating that ASD–associated patterns are not merely developmental artifacts. However, the magnitude and direction of taxa-level changes varied with age, underscoring the need for age–stratified analyses in paediatric microbiome studies.

Early–life ASD toddlers exhibited depletion of beneficial taxa (*Bifidobacterium*, *Blautia*), children showed fermentative dysbiosis (*Collinsella*, *Dysosmobacter*, *Ruthenibacterium*), and adolescents exhibited mature dysbiosis dominated by the reduction of mucin degraders (*Akkermansia*), anaerobic fermenters (*Alistipes*, *Phocaeicola*), and taxa associated with chronic inflammation or altered SCFA metabolism. Overall, ASD-associated dysbiosis persisted across childhood but appeared less taxonomically pronounced with age, possibly reflecting dietary diversification or microbiome convergence during adolescence.

ML analyses corroborated these findings. ASD toddlers’ microbiota showed the strongest predictive signal, gradually declining in children and adolescents. Key taxa varied by age but consistently included *Faecalibacterium*, *Bacteroides*_H, *Phocaeicola*, *Akkermansia*, and *Roseburia*. ASD appears associated with an altered microbiota maturation trajectory, with early-life dysbiosis persisting rather than shifting toward neurotypical age-related patterns. This may have mechanistic relevance, as early childhood is critical for gut–brain signalling, immune education, and metabolic programming (Dera et al., 2025; Laue et al., 2022; Voreades et al., 2014).

Network analyses showed age–dependent shifts in microbial interactions, with toddlers displaying smaller, less connected networks than children and adolescents, consistent with microbiome maturation and increasing network density (Zou et al., 2026). These dynamics may influence neuroimmune and metabolic pathways implicated in ASD (Cryan et al., 2019).

Metabolomic profiling revealed distinct alterations in faecal VOCs and SCFAs in ASD, indicating a metabolically divergent gut environment. Benzenamine, N-ethyl-, 5-Hepten-2-one, 6-methyl-, and methyl isobutyl ketone were consistently significant, suggesting biomarker potential. Elevated methyl isobutyl ketone and 6-methyl-5-hepten-2-one reflect altered fermentation, while N-ethyl benzenamine likely derives from amino acid metabolism (Diether and Willing, 2019; Vernocchi et al., 2023). Notably, 6-methyl-5-hepten-2-one correlated positively with CBCL-EXT, and methyl isobutyl ketone negatively with CBCL-INT, supporting links between microbial metabolites and neurobehavior (Cryan et al., 2019; Sharon et al., 2019).

Other VOCs, including methional, 1-Heptanol, and 2-Butanone, were reduced in ASD, reflecting altered sulfur amino acid, FA, and carbohydrate metabolism (Garner et al., 2007; Indika et al., 2021; Vernocchi et al., 2023).

Methyl salicylate correlated positively with *Mediterraneanbacter* and *Moraxella*, and pentanoic acid, ethyl ester with SCFA-producing taxa (*Agathobacter*, *Butyricicoccus*, *Megasphaera*, *Eubacterium*), indicating distinct metabolic configurations within the ASD gut microbiota (Cryan et al., 2019; Vernocchi et al., 2023; Zierer et al., 2018). Moreover, methyl salicylate was also correlated with CBCL-TOT and CBCL-EXT. In piglets, methyl salicylate plus tributyrin improved intestinal morphology, microbiota composition, and growth performance, increasing villus height, Firmicutes/Bacteroides ratio, amino acid metabolism, and arginine biosynthesis, while reducing *Campylobacter* abundance (Wei et al., 2021; Zhang et al., 2020).

Age–specific VOC profiling showed marked alterations in toddlers, with elevated aromatic and indole compounds (benzaldehyde, indole, p-cresol, benzothiazole), consistent with sparser microbial networks and an immature dysbiotic microbiota. Differences decreased in older groups as networks became denser and more interconnected.

Indole and its derivative 3-methylindole (skatole) are microbial metabolites from tryptophan catabolism (Li et al., 2021). Studies report elevated indole derivatives in ASD urine and feces, reflecting altered microbial tryptophanase activity in taxa such as *Escherichia coli*, *Proteus vulgaris*, *Paracolobactrum coliform*, *Achromobacter liquefaciens*, and *Bacteroides* spp. (Gevi et al., 2016; Keszthelyi et al., 2009). While altered tryptophan metabolism correlates with ASD symptoms and gut changes, causality remains unproven (Aziz-Zadeh et al., 2025). Elevated indole and 3-methylindole in ASD toddlers likely indicate early-life microbial immaturity, supported by network analysis.

SCFA profiling showed increased acetic and propanoic acids but reduced butanoic and pentanoic acids; elevated propanoic acid can modulate behaviour and neuroinflammation (Meeking et al., 2020). Age-stratified analyses revealed that toddlers had broadly elevated SCFAs, children exhibited selective acetic acid elevation, and adolescents showed a pattern resembling the overall cohort.

D-AA analysis revealed a consistent reduction of D-aspartate and D-alanine in ASD across all ages. D-AAs are increasingly recognized as important microbiota-derived signalling molecules, with D- aspartate acting as an endogenous modulator of N-Methyl-D-Aspartate- (NMDA-) receptor–mediated neurotransmission and neurodevelopmental processes, while D-alanine contributes to neurotransmission, immune and epithelial regulation (Errico et al., 2020, 2018; Lee et al., 2022, 2020; Usiello et al., 2020). Correlations with specific taxa (i.e., *Alistipes*, *Ruthenibacterium*, *Gemmiger*, *Lawsonibacter*, *Streptococcus*, *Bifidobacterium*, and *Eubacterium*) suggest functional reprogramming of microbial D-AA metabolism in ASD, potentially impacting gut–brain and neuroimmune signalling during critical developmental windows.

Interestingly, experimental studies in environmental rat models of ASD have shown that D-aspartate alterations were primarily detectable in the brain, where prenatal inflammation or valproate exposure induces region-specific accumulation during adolescence, while peripheral compartments such as plasma and feces remained unchanged (Di Maio et al., 2025), suggesting a degree of compartmentalization of D-AA regulation between the central nervous system and the intestinal environment. The apparent discrepancy with our human findings may reflect species–related differences in gut microbiota composition and metabolic activity, which can influence microbial D-AA production and degradation within the intestinal ecosystem, thereby modulating faecal D-aspartate pool independently of central levels. However, the role of microbiota–associated D-aspartate in the gut–brain axis and its potential involvement in ASD remains largely unexplored. Therefore, future studies will be required to determine whether faecal D-aspartate changes observed in ASD patients are linked to central D-aspartate dynamics or reflect microbiota–specific metabolic alterations associated with the disorder.

## Conclusion

In summary, our integrated analysis of bacterial taxa, VOCs, SCFAs, and D-AAs reveals that ASD is associated with early-life gut microbial dysbiosis and altered metabolic outputs, most pronounced in toddlers, coinciding with smaller, sparser microbial networks and elevated aromatic and indole-derived metabolites, a critical window for microbiome–host interactions. Dysbiosis persists across childhood but shows age–dependent shifts toward more mature microbial configurations and attenuated metabolite differences. Functional alterations, including disrupted SCFA profiles and reduced D-AAs, underscore perturbations in fermentation, AA metabolism, and gut–brain signalling. Clinically, these findings highlight the potential for early microbiome–targeted interventions, such as dietary modulation, prebiotics, or probiotics, to restore microbial balance and metabolic function. Translationally, the identified microbial metabolite signatures and selective D-AA variations may serve as potential biomarkers for ASD diagnosis, prognosis, or personalized therapeutic strategies, emphasizing the gut microbiota as a modifiable contributor to neurodevelopmental outcomes.

## Supporting information

Supplementary meterials

## Acknowledgments

This work was supported by the Italian Ministry of Health with “Current Research funds” and with Network IDEA (Project titled “Il microbiota umano nella medicina dei sistemi applicata alla diagnosi e alla terapia in pediatria”) and by Italian Ministry of University and Research with PRIN PNRR 2022, financed by the European Union – Next Generation EU (P2022ZEMZF, Project titled “D-aspartate metabolism dysfunction as an emerging biochemical signature in neurodevelopmental psychiatric disorders”).

## Conflicts of Interest

The authors declare no conflicts of interest.

## Data Availability Statement

The datasets generated and analyzed in this study are available from the corresponding author upon reasonable request, in accordance with ethical and privacy regulations.

## Supplementary Information

The Supplementary Materials include 17 figures providing extended methodological and analytical details supporting the main findings. Figure S1 outlines the study design and multi-omics workflow in ASD and control paediatric cohorts stratified by age. Figures S2–S6 report additional gut microbiota analyses, including diversity metrics, differential abundance, machine learning classification, and age-stratified compositional changes. Figures S7–S8 present extended metabolomic results, including VOC profiling, multivariate modelling, and classification performance. Figures S9–S13 provide associations between metabolites and behavioural, clinical, and SCFA-related measures, including regression and age-stratified analyses. Figures S14–S17 report comprehensive correlation analyses integrating bacterial taxa, amino acids, and metabolites across age groups.

## Notes

### Competing Interest Statement

The authors have declared no competing interest.

### Author Declarations

Ethics committee/IRB of Bambino Gesu Children s Hospital gave ethical approval for this work

## References

American Psychiatric Association, 2013. Diagnostic and statistical manual of mental disorders: DSM-5, 5th ed. ed. American psychiatric association, Washington.

Aziz-Zadeh, L., Ringold, S.M., Jayashankar, A., Kilroy, E., Butera, C., Jacobs, J.P., Tanartkit, S., Mahurkar-Joshi, S., Bhatt, R.R., Dapretto, M., Labus, J.S., Mayer, E.A., 2025. Relationships between brain activity, tryptophan-related gut metabolites, and autism symptomatology. Nat Commun 16, 3465. 10.1038/s41467-025-58459-1

Bäckhed, F., Roswall, J., Peng, Y., Feng, Q., Jia, H., Kovatcheva-Datchary, P., Li, Y., Xia, Y., Xie, H., Zhong, H., Khan, M.T., Zhang, J., Li, J., Xiao, L., Al-Aama, J., Zhang, D., Lee, Y.S., Kotowska, D., Colding, C., Tremaroli, V., Yin, Y., Bergman, S., Xu, X., Madsen, L., Kristiansen, K., Dahlgren, J., Wang, J., 2015. Dynamics and Stabilization of the Human Gut Microbiome during the First Year of Life. Cell Host & Microbe 17, 690–703. 10.1016/j.chom.2015.04.004

Belenguer, A., Duncan, S.H., Calder, A.G., Holtrop, G., Louis, P., Lobley, G.E., Flint, H.J., 2006. Two Routes of Metabolic Cross-Feeding between *Bifidobacterium adolescentis* and Butyrate-Producing Anaerobes from the Human Gut. Appl Environ Microbiol 72, 3593–3599. 10.1128/AEM.72.5.3593-3599.2006

Bolyen, E., Rideout, J.R., Dillon, M.R., Bokulich, N.A., Abnet, C.C., Al-Ghalith, G.A., Alexander, H., Alm, E.J., Arumugam, M., Asnicar, F., Bai, Y., Bisanz, J.E., Bittinger, K., Brejnrod, A., Brislawn, C.J., Brown, C.T., Callahan, B.J., Caraballo-Rodríguez, A.M., Chase, J., Cope, E.K., Da Silva, R., Diener, C., Dorrestein, P.C., Douglas, G.M., Durall, D.M., Duvallet, C., Edwardson, C.F., Ernst, M., Estaki, M., Fouquier, J., Gauglitz, J.M., Gibbons, S.M., Gibson, D.L., Gonzalez, A., Gorlick, K., Guo, J., Hillmann, B., Holmes, S., Holste, H., Huttenhower, C., Huttley, G.A., Janssen, S., Jarmusch, A.K., Jiang, L., Kaehler, B.D., Kang, K.B., Keefe, C.R., Keim, P., Kelley, S.T., Knights, D., Koester, I., Kosciolek, T., Kreps, J., Langille, M.G.I., Lee, J., Ley, R., Liu, Y.-X., Loftfield, E., Lozupone, C., Maher, M., Marotz, C., Martin, B.D., McDonald, D., McIver, L.J., Melnik, A.V., Metcalf, J.L., Morgan, S.C., Morton, J.T., Naimey, A.T., Navas-Molina, J.A., Nothias, L.F., Orchanian, S.B., Pearson, T., Peoples, S.L., Petras, D., Preuss, M.L., Pruesse, E., Rasmussen, L.B., Rivers, A., Robeson, M.S., Rosenthal, P., Segata, N., Shaffer, M., Shiffer, A., Sinha, R., Song, S.J., Spear, J.R., Swafford, A.D., Thompson, L.R., Torres, P.J., Trinh, P., Tripathi, A., Turnbaugh, P.J., Ul-Hasan, S., Van Der Hooft, J.J.J., Vargas, F., Vázquez-Baeza, Y., Vogtmann, E., Von Hippel, M., Walters, W., Wan, Y., Wang, M., Warren, J., Weber, K.C., Williamson, C.H.D., Willis, A.D., Xu, Z.Z., Zaneveld, J.R., Zhang, Y., Zhu, Q., Knight, R., Caporaso, J.G., 2019. Author Correction: Reproducible, interactive, scalable and extensible microbiome data science using QIIME 2. Nat Biotechnol 37, 1091–1091. 10.1038/s41587-019-0252-6

Callahan, B.J., McMurdie, P.J., Rosen, M.J., Han, A.W., Johnson, A.J.A., Holmes, S.P., 2016. DADA2: High-resolution sample inference from Illumina amplicon data. Nat Methods 13, 581–583. 10.1038/nmeth.3869

Caputi, V., Hill, L., Figueiredo, M., Popov, J., Hartung, E., Margolis, K.G., Baskaran, K., Joharapurkar, P., Moshkovich, M., Pai, N., 2024. Functional contribution of the intestinal microbiome in autism spectrum disorder, attention deficit hyperactivity disorder, and Rett syndrome: a systematic review of pediatric and adult studies. Front. Neurosci. 18, 1341656. 10.3389/fnins.2024.1341656

Chamtouri, M., Gaddour, N., Merghni, A., Mastouri, M., Arboleya, S., De Los Reyes-Gavilán, C.G., 2023. Age and severity-dependent gut microbiota alterations in Tunisian children with autism spectrum disorder. Sci Rep 13, 18218. 10.1038/s41598-023-45534-0

Chen, Q., Xu, J., Yang, J., Qin, X., Fan, J., Ke, H., Yang, Z., Zheng, W., Li, X., Huang, L., Ning, W., 2025. Gut microbiota analysis in children with autism spectrum disorder and their family members. Sci Rep 15, 44282. 10.1038/s41598-025-27822-z

Cryan, J.F., O’Riordan, K.J., Cowan, C.S.M., Sandhu, K.V., Bastiaanssen, T.F.S., Boehme, M., Codagnone, M.G., Cussotto, S., Fulling, C., Golubeva, A.V., Guzzetta, K.E., Jaggar, M., Long-Smith, C.M., Lyte, J.M., Martin, J.A., Molinero-Perez, A., Moloney, G., Morelli, E., Morillas, E., O’Connor, R., Cruz-Pereira, J.S., Peterson, V.L., Rea, K., Ritz, N.L., Sherwin, E., Spichak, S., Teichman, E.M., Van De Wouw, M., Ventura-Silva, A.P., Wallace-Fitzsimons, S.E., Hyland, N., Clarke, G., Dinan, T.G., 2019. The Microbiota-Gut-Brain Axis. Physiological Reviews 99, 1877–2013. 10.1152/physrev.00018.2018

Dalile, B., Van Oudenhove, L., Vervliet, B., Verbeke, K., 2019. The role of short-chain fatty acids in microbiota–gut–brain communication. Nat Rev Gastroenterol Hepatol 16, 461–478. 10.1038/s41575-019-0157-3

Dera, N., Kosińska-Kaczyńska, K., Żeber-Lubecka, N., Brawura-Biskupski-Samaha, R., Massalska, D., Szymusik, I., Dera, K., Ciebiera, M., 2025. Impact of Early-Life Microbiota on Immune System Development and Allergic Disorders. Biomedicines 13, 121. 10.3390/biomedicines13010121

Di Maio, A., Yahyavi, I., Buzzelli, V., Motta, Z., Ascone, F., Putignani, L., Usiello, A., Pollegioni, L., Trezza, V., Errico, F., 2025. Prenatal Exposure to Lipopolysaccharide or Valproate Leads to Abnormal Accumulation of the NMDA Receptor Agonist D-Aspartate in the Adolescent Rat Brain. Journal of Neurochemistry 169, e70095. 10.1111/jnc.70095

Diether, N.E., Willing, B.P., 2019. Microbial Fermentation of Dietary Protein: An Important Factor in Diet–Microbe–Host Interaction. Microorganisms 7, 19. 10.3390/microorganisms7010019

Errico, F., Cuomo, M., Canu, N., Caputo, V., Usiello, A., 2020. New insights on the influence of free d-aspartate metabolism in the mammalian brain during prenatal and postnatal life. Biochimica et Biophysica Acta (BBA) - Proteins and Proteomics 1868, 140471. 10.1016/j.bbapap.2020.140471

Errico, F., Nuzzo, T., Carella, M., Bertolino, A., Usiello, A., 2018. The Emerging Role of Altered d-Aspartate Metabolism in Schizophrenia: New Insights From Preclinical Models and Human Studies. Front. Psychiatry 9, 559. 10.3389/fpsyt.2018.00559

Errico, F., Russo, R., Carrillo, F., Nuzzo, T., di Vito, R., Canonico, E., Pedone, P.V., Di Cunto, F., Esposito, T., Usiello, A., Chambery, A., 2025. Free D-aspartate modulates the expression of proteins linked to schizophrenia and autism spectrum disorder during early postnatal life. Cell Mol Life Sci 83, 3. 10.1007/s00018-025-05938-9

Esler, A.N., Bal, V.H., Guthrie, W., Wetherby, A., Weismer, S.E., Lord, C., 2015. The autism diagnostic observation schedule, toddler module: Standardized severity scores. Journal of autism and developmental disorders 45, 2704–2720.

Flint, H.J., Scott, K.P., Duncan, S.H., Louis, P., Forano, E., 2012. Microbial degradation of complex carbohydrates in the gut. Gut Microbes 3, 289–306. 10.4161/gmic.19897

Garner, C.E., Smith, S., De Lacy Costello, B., White, P., Spencer, R., Probert, C.S.J., Ratcliffem, N.M., 2007. Volatile organic compounds from feces and their potential for diagnosis of gastrointestinal disease. The FASEB Journal 21, 1675–1688. 10.1096/fj.06-6927com

Gevi, F., Zolla, L., Gabriele, S., Persico, A.M., 2016. Urinary metabolomics of young Italian autistic children supports abnormal tryptophan and purine metabolism. Molecular Autism 7, 47. 10.1186/s13229-016-0109-5

Gonda, Y., Matsuda, A., Adachi, K., Ishii, C., Suzuki, M., Osaki, A., Mita, M., Nishizaki, N., Ohtomo, Y., Shimizu, T., Yasui, M., Hamase, K., Sasabe, J., 2023. Mammals sustain amino acid homochirality against chiral conversion by symbiotic microbes. Proc. Natl. Acad. Sci. U.S.A. 120, e2300817120. 10.1073/pnas.2300817120

Gotham, K., Pickles, A., Lord, C., 2009. Standardizing ADOS Scores for a Measure of Severity in Autism Spectrum Disorders. J Autism Dev Disord 39, 693–705. 10.1007/s10803-008-0674-3

Hodges, H., Fealko, C., Soares, N., 2020. Autism spectrum disorder: definition, epidemiology, causes, and clinical evaluation. Transl Pediatr 9, S55–S65. 10.21037/tp.2019.09.09

Hus, V., Lord, C., 2014. The autism diagnostic observation schedule, module 4: revised algorithm and standardized severity scores. Journal of autism and developmental disorders 44, 1996–2012.

Iljazovic, A., Roy, U., Gálvez, E.J.C., Lesker, T.R., Zhao, B., Gronow, A., Amend, L., Will, S.E., Hofmann, J.D., Pils, M.C., Schmidt-Hohagen, K., Neumann-Schaal, M., Strowig, T., 2021. Perturbation of the gut microbiome by Prevotella spp. enhances host susceptibility to mucosal inflammation. Mucosal Immunology 14, 113–124. 10.1038/s41385-020-0296-4

Indika, N.-L.R., Deutz, N.E.P., Engelen, M.P.K.J., Peiris, H., Wijetunge, S., Perera, R., 2021. Sulfur amino acid metabolism and related metabotypes of autism spectrum disorder: A review of biochemical evidence for a hypothesis. Biochimie 184, 143–157. 10.1016/j.biochi.2021.02.018

Kadiyska, T., Vassilev, D., Tourtourikov, I., Ciurinskiene, S., Madzharova, D., Savcheva, M., Stoynev, N., Mileva-Popova, R., Tafradjiiska-Hadjiolova, R., Mitev, V., 2025. Age-Dependent Gut Microbiome Dysbiosis in Autism Spectrum Disorder and the Role of Key Bacterial Ratios. Nutrients 17, 1775. 10.3390/nu17111775

Keszthelyi, D., Troost, F., Masclee, A., 2009. Understanding the role of tryptophan and serotonin metabolism in gastrointestinal function. Neurogastroenterology & Motility 21, 1239–1249.

Koh, A., De Vadder, F., Kovatcheva-Datchary, P., Bäckhed, F., 2016. From Dietary Fiber to Host Physiology: Short-Chain Fatty Acids as Key Bacterial Metabolites. Cell 165, 1332–1345. 10.1016/j.cell.2016.05.041

Korteniemi, J., Karlsson, L., Aatsinki, A., 2024. Systematic Review: Autism Spectrum Disorder and the Gut Microbiota. FOC 22, 242–251. 10.1176/appi.focus.24022008

La Rosa, P.S., Warner, B.B., Zhou, Y., Weinstock, G.M., Sodergren, E., Hall-Moore, C.M., Stevens, H.J., Bennett, W.E., Shaikh, N., Linneman, L.A., Hoffmann, J.A., Hamvas, A., Deych, E., Shands, B.A., Shannon, W.D., Tarr, P.I., 2014. Patterned progression of bacterial populations in the premature infant gut. Proc. Natl. Acad. Sci. U.S.A. 111, 12522–12527. 10.1073/pnas.1409497111

Laue, H.E., Coker, M.O., Madan, J.C., 2022. The Developing Microbiome From Birth to 3 Years: The Gut-Brain Axis and Neurodevelopmental Outcomes. Front. Pediatr. 10, 815885. 10.3389/fped.2022.815885

Lee, C.J., Qiu, T.A., Hong, Z., Zhang, Z., Min, Y., Zhang, L., Dai, L., Zhao, H., Si, T., Sweedler, J.V., 2022. Profiling of d-alanine production by the microbial isolates of rat gut microbiota. FASEB J 36, e22446. 10.1096/fj.202101595R

Lee, C.J., Qiu, T.A., Sweedler, J.V., 2020. d-Alanine: Distribution, origin, physiological relevance, and implications in disease. Biochim Biophys Acta Proteins Proteom 1868, 140482. 10.1016/j.bbapap.2020.140482

Li, X., Zhang, B., Hu, Y., Zhao, Y., 2021. New Insights Into Gut-Bacteria-Derived Indole and Its Derivatives in Intestinal and Liver Diseases. Front. Pharmacol. 12, 769501. 10.3389/fphar.2021.769501

Louis, P., Flint, H.J., 2017. Formation of propionate and butyrate by the human colonic microbiota. Environmental Microbiology 19, 29–41. 10.1111/1462-2920.13589

Maenner, M.J., Warren, Z., Williams, A.R., Amoakohene, E., Bakian, A.V., Bilder, D.A., Durkin, M.S., Fitzgerald, R.T., Furnier, S.M., Hughes, M.M., Ladd-Acosta, C.M., McArthur, D., Pas, E.T., Salinas, A., Vehorn, A., Williams, S., Esler, A., Grzybowski, A., Hall-Lande, J., Nguyen, R.H.N., Pierce, K., Zahorodny, W., Hudson, A., Hallas, L., Mancilla, K.C., Patrick, M., Shenouda, J., Sidwell, K., DiRienzo, M., Gutierrez, J., Spivey, M.H., Lopez, M., Pettygrove, S., Schwenk, Y.D., Washington, A., Shaw, K.A., 2023. Prevalence and Characteristics of Autism Spectrum Disorder Among Children Aged 8 Years — Autism and Developmental Disabilities Monitoring Network, 11 Sites, United States, 2020. MMWR Surveill. Summ. 72, 1–14. 10.15585/mmwr.ss7202a1

Marangelo, C., Vernocchi, P., Del Chierico, F., Scanu, M., Marsiglia, R., Petrolo, E., Fucà, E., Guerrera, S., Valeri, G., Vicari, S., Putignani, L., 2024. Stratification of Gut Microbiota Profiling Based on Autism Neuropsychological Assessments. Microorganisms 12, 2041. 10.3390/microorganisms12102041

McDonald, D., Jiang, Y., Balaban, M., Cantrell, K., Zhu, Q., Gonzalez, A., Morton, J.T., Nicolaou, G., Parks, D.H., Karst, S.M., Albertsen, M., Hugenholtz, P., DeSantis, T., Song, S.J., Bartko, A., Havulinna, A.S., Jousilahti, P., Cheng, S., Inouye, M., Niiranen, T., Jain, M., Salomaa, V., Lahti, L., Mirarab, S., Knight, R., 2024. Author Correction: Greengenes2 unifies microbial data in a single reference tree. Nat Biotechnol 42, 813–813. 10.1038/s41587-023-02026-w

Meeking, M.M., MacFabe, D.F., Mepham, J.R., Foley, K.A., Tichenoff, L.J., Boon, F.H., Kavaliers, M., Ossenkopp, K.-P., 2020. Propionic acid induced behavioural effects of relevance to autism spectrum disorder evaluated in the hole board test with rats. Progress in Neuro- Psychopharmacology and Biological Psychiatry 97, 109794. 10.1016/j.pnpbp.2019.109794

Novau-Ferré, N., Papandreou, C., Rojo-Marticella, M., Canals-Sans, J., Bulló, M., 2025. Gut microbiome differences in children with Attention Deficit Hyperactivity Disorder and Autism Spectrum Disorder and effects of probiotic supplementation: A randomized controlled trial. Research in Developmental Disabilities 161, 105003. 10.1016/j.ridd.2025.105003

Nuzzo, T., Sekine, M., Punzo, D., Miroballo, M., Katane, M., Saitoh, Y., Galbusera, A., Pasqualetti, M., Errico, F., Gozzi, A., Mothet, J.-P., Homma, H., Usiello, A., 2020. Dysfunctional d-aspartate metabolism in BTBR mouse model of idiopathic autism. Biochim Biophys Acta Proteins Proteom 1868, 140531. 10.1016/j.bbapap.2020.140531

Parada Venegas, D., De La Fuente, M.K., Landskron, G., González, M.J., Quera, R., Dijkstra, G., Harmsen, H.J.M., Faber, K.N., Hermoso, M.A., 2019. Short Chain Fatty Acids (SCFAs)-Mediated Gut Epithelial and Immune Regulation and Its Relevance for Inflammatory Bowel Diseases. Front. Immunol. 10, 277. 10.3389/fimmu.2019.00277

Puricelli, C., Rolla, R., Gigliotti, L., Boggio, E., Beltrami, E., Dianzani, U., Keller, R., 2022. The Gut-Brain-Immune Axis in Autism Spectrum Disorders: A State-of-Art Report. Front. Psychiatry 12, 755171. 10.3389/fpsyt.2021.755171

Scanu, M., Del Chierico, F., Marsiglia, R., Toto, F., Guerrera, S., Valeri, G., Vicari, S., Putignani, L., 2024. Correction of Batch Effect in Gut Microbiota Profiling of ASD Cohorts from Different Geographical Origins. Biomedicines 12, 2350. 10.3390/biomedicines12102350

Segata, N., Izard, J., Waldron, L., Gevers, D., Miropolsky, L., Garrett, W.S., Huttenhower, C., 2011. Metagenomic biomarker discovery and explanation. Genome Biol 12, R60. 10.1186/gb-2011-12-6-r60

Serra, M., Di Maio, A., Bassareo, V., Nuzzo, T., Errico, F., Servillo, F., Capasso, M., Parekh, P., Li, Q., Thiolat, M.-L., Bezard, E., Calabresi, P., Sulzer, D., Carta, M., Morelli, M., Usiello, A., 2023. Perturbation of serine enantiomers homeostasis in the striatum of MPTP-lesioned monkeys and mice reflects the extent of dopaminergic midbrain degeneration. Neurobiology of Disease 184, 106226. 10.1016/j.nbd.2023.106226

Sharon, G., Cruz, N.J., Kang, D.-W., Gandal, M.J., Wang, B., Kim, Y.-M., Zink, E.M., Casey, C.P., Taylor, B.C., Lane, C.J., Bramer, L.M., Isern, N.G., Hoyt, D.W., Noecker, C., Sweredoski, M.J., Moradian, A., Borenstein, E., Jansson, J.K., Knight, R., Metz, T.O., Lois, C., Geschwind, D.H., Krajmalnik-Brown, R., Mazmanian, S.K., 2019. Human Gut Microbiota from Autism Spectrum Disorder Promote Behavioral Symptoms in Mice. Cell 177, 1600–1618.e17. 10.1016/j.cell.2019.05.004

Silva, Y.P., Bernardi, A., Frozza, R.L., 2020. The Role of Short-Chain Fatty Acids From Gut Microbiota in Gut-Brain Communication. Front. Endocrinol. 11, 25. 10.3389/fendo.2020.00025

Socała, K., Doboszewska, U., Szopa, A., Serefko, A., Włodarczyk, M., Zielińska, A., Poleszak, E., Fichna, J., Wlaź, P., 2021. The role of microbiota-gut-brain axis in neuropsychiatric and neurological disorders. Pharmacological Research 172, 105840. 10.1016/j.phrs.2021.105840

Stewart, C.J., Ajami, N.J., O’Brien, J.L., Hutchinson, D.S., Smith, D.P., Wong, M.C., Ross, M.C., Lloyd, R.E., Doddapaneni, H., Metcalf, G.A., Muzny, D., Gibbs, R.A., Vatanen, T., Huttenhower, C., Xavier, R.J., Rewers, M., Hagopian, W., Toppari, J., Ziegler, A.-G., She, J.-X., Akolkar, B., Lernmark, A., Hyoty, H., Vehik, K., Krischer, J.P., Petrosino, J.F., 2018. Temporal development of the gut microbiome in early childhood from the TEDDY study. Nature 562, 583–588. 10.1038/s41586-018-0617-x

Toscano De Oliveira, M., Borges Scriboni Gonzalez, M.A., Rosetto Boiate, J.V., Mesa, V., Gonçalves, D.H., Gisse Pinto, M., Ramos Da Silva Pinto, C., Costa Gomes Filho, J.E., Sivieri, K., 2025. Altered gut microbiota composition and feeding behaviours in children with Autism Spectrum Disorder: a comparative pilot study. Annals of Human Biology 52, 2535430. 10.1080/03014460.2025.2535430

Turroni, F., Milani, C., Duranti, S., Ferrario, C., Lugli, G.A., Mancabelli, L., Van Sinderen, D., Ventura, M., 2018. Bifidobacteria and the infant gut: an example of co-evolution and natural selection. Cell. Mol. Life Sci. 75, 103–118. 10.1007/s00018-017-2672-0

Usiello, A., Di Fiore, M.M., De Rosa, A., Falvo, S., Errico, F., Santillo, A., Nuzzo, T., Chieffi Baccari, G., 2020. New Evidence on the Role of D-Aspartate Metabolism in Regulating Brain and Endocrine System Physiology: From Preclinical Observations to Clinical Applications. Int J Mol Sci 21, 8718. 10.3390/ijms21228718

Valencia-Buitrago, M., Oliveira-Carvalho, R.D., Cardoso, V., Triviño-Valencia, J., Salamanca-Duque, L.M., Martínez-Díaz, V., Zabaleta, J., Galeano-Vanegas, N.F., Naranjo-Galvis, C.A., 2025. Metagenomic Characterization of Gut Microbiota in Children with Autism Spectrum Disorder: Microbial Signatures and Modulation by Anti-Inflammatory Diet and Probiotics. Pharmaceuticals 18, 1376. 10.3390/ph18091376

Vernocchi, P., Marangelo, C., Guerrera, S., Del Chierico, F., Guarrasi, V., Gardini, S., Conte, F., Paci, P., Ianiro, G., Gasbarrini, A., Vicari, S., Putignani, L., 2023. Gut microbiota functional profiling in autism spectrum disorders: bacterial VOCs and related metabolic pathways acting as disease biomarkers and predictors. Front. Microbiol. 14, 1287350. 10.3389/fmicb.2023.1287350

Vernocchi, P., Marini, F., Capuani, G., Tomassini, A., Conta, G., Del Chierico, F., Malattia, C., De Benedetti, F., Martini, A., Dallapiccola, B., Van Dijkhuizen, E.H.P., Miccheli, A., Putignani, L., 2020. Fused Omics Data Models Reveal Gut Microbiome Signatures Specific of Inactive Stage of Juvenile Idiopathic Arthritis in Pediatric Patients. Microorganisms 8, 1540. 10.3390/microorganisms8101540

Vernocchi, P., Ristori, M.V., Guerrera, S., Guarrasi, V., Conte, F., Russo, A., Lupi, E., Albitar-Nehme, S., Gardini, S., Paci, P., Ianiro, G., Vicari, S., Gasbarrini, A., Putignani, L., 2022. Gut Microbiota Ecology and Inferred Functions in Children With ASD Compared to Neurotypical Subjects. Front. Microbiol. 13, 871086. 10.3389/fmicb.2022.871086

Voreades, N., Kozil, A., Weir, T.L., 2014. Diet and the development of the human intestinal microbiome. Front. Microbiol. 5. 10.3389/fmicb.2014.00494

Wei, Y., Mao, J., Liu, J., Zhang, Y., Deng, Z., Lv, J., He, M., Liu, J., Wang, H., 2021. Encapsulated mixture of methyl salicylate and tributyrin modulates intestinal microbiota and improves growth performance of weaned piglets. Microorganisms 9. 10.3390/microorganisms9061342

Xiang, F., Zhang, M., Wei, X., Chang, J., 2025. Gut microbiota composition and phylogenetic analysis in autism spectrum disorder: a comparative study. Front. Psychiatry 16, 1609638. 10.3389/fpsyt.2025.1609638

Yang, H., Wang, A., Yang, J., Luo, R., Yang, Y., 2025. Alterations in gut microbiota composition in neurodevelopmental disorders: a systematic review and meta-analysis. Front. Microbiol. 16, 1650212. 10.3389/fmicb.2025.1650212

Yatsunenko, T., Rey, F.E., Manary, M.J., Trehan, I., Dominguez-Bello, M.G., Contreras, M., Magris, M., Hidalgo, G., Baldassano, R.N., Anokhin, A.P., Heath, A.C., Warner, B., Reeder, J., Kuczynski, J., Caporaso, J.G., Lozupone, C.A., Lauber, C., Clemente, J.C., Knights, D., Knight, R., Gordon, J.I., 2012. Human gut microbiome viewed across age and geography. Nature 486, 222–227. 10.1038/nature11053

Zhang, W.-X., Zhang, Y., Zhang, X.-W., Deng, Z.-X., Liu, J.-X., He, M.-L., Wang, H.-F., 2020. Effects of dietary supplementation with combination of tributyrin and essential oil on gut health and microbiota of weaned piglets. Animals 10. 10.3390/ani10020180

Zierer, J., Jackson, M.A., Kastenmüller, G., Mangino, M., Long, T., Telenti, A., Mohney, R.P., Small, K.S., Bell, J.T., Steves, C.J., Valdes, A.M., Spector, T.D., Menni, C., 2018. The fecal metabolome as a functional readout of the gut microbiome. Nat Genet 50, 790–795. 10.1038/s41588-018-0135-7

Zou, B., Huo, Q., Zhou, X., Lv, Y., Li, G., Fu, G., Shen, H., Shu, S., 2026. Characteristics and longitudinal stability of Gut Microbiota in healthy individuals across different age groups. Current Research in Microbial Sciences 10, 100512. 10.1016/j.crmicr.2025.100512

