## Supplementary meterials for "Developmental Trajectories of Gut Microbiota and Microbial Metabolites in Children with Autism Spectrum Disorder"

### Supplementary materials

#### Supplementary figures

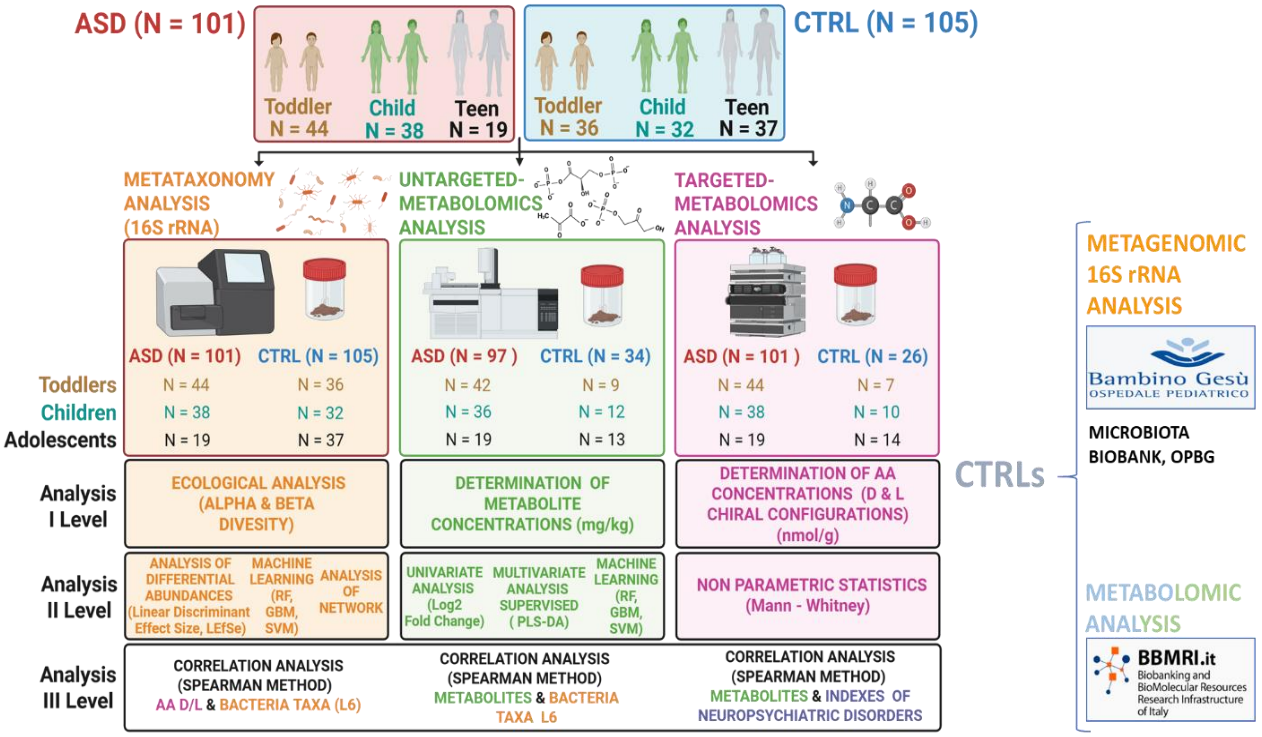


**Figure S1.** Study design and multi-omics analyses workflow of the paediatric ASD and neurotypical cohorts stratified by age group. The study enrolled 101 children with autism spectrum disorder (ASD) and 105 neurotypical controls (CTRL), divided into three age groups: toddlers (ASD N=44, CTRL N=36), children (ASD N=38, CTRL N=32), and adolescents (ASD N=19, CTRL N=37). Three main omics analyses were performed on faecal samples: metataxonomy analysis (16S rRNA sequencing); untargeted metabolomics analysis and targeted metabolomics analysis. Data analyses were organized at three levels: I) ecological diversity (alpha and beta diversity) for microbiome data and concentration measurements for metabolites and amino acids; II) differential abundance analysis, machine learning (Random Forest, Gradient Boosting Machine, Support Vector Machine), and network analysis for microbiome and metabolomics data. Statistical tests included linear discriminant effect size (LEfSe) and Mann-Whitney nonparametric tests; III) correlation analyses using Spearman's method between amino acids and bacterial taxa, metabolites and bacterial taxa, and metabolites and neuropsychiatric disorder indexes.

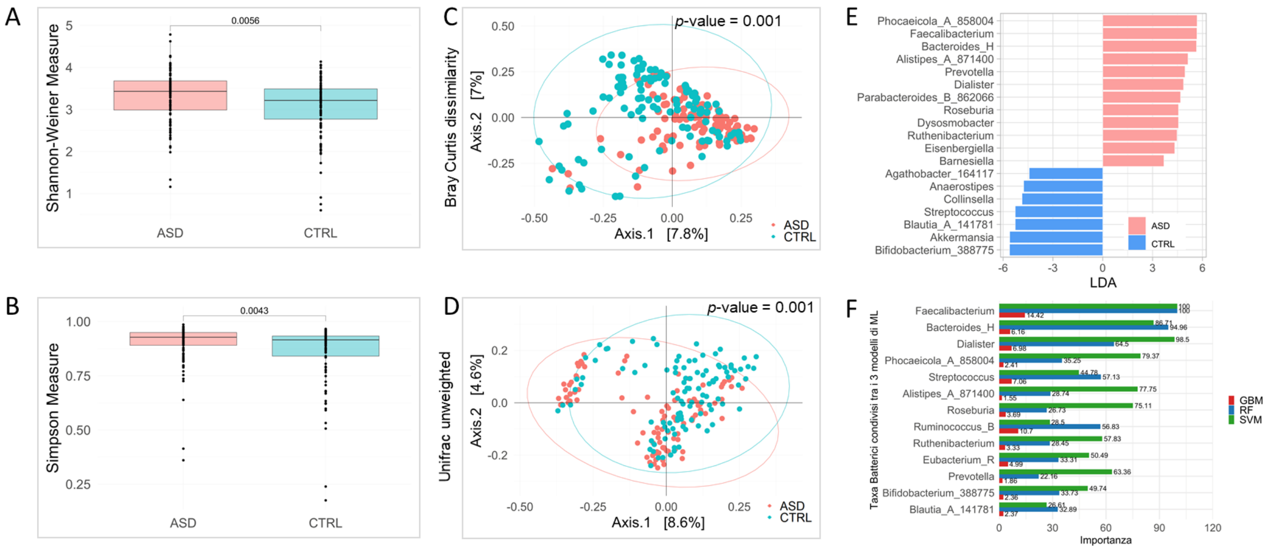


**Figure S2: Gut microbiota ecology in ASD vs CTRL. (A)** Boxplots comparing Shannon diversity in ASD and CTRL groups. **(B)** Boxplots comparing Simpson index diversity in ASD and CTRL groups. Wilcoxon rank-sum p-values are reported. (**C)** Bray–Curtis PCoA plot of ASD vs CTRL. (**D)** Unweighted UniFrac PCoA plot of ASD vs CTRL. PERMANOVA p values are shown. **(E)** Differential taxa (LEfSe) LDA score plot showing taxa discriminating between ASD and CTRL at genus level. **(F)** Machine Learning (ML) classifier performance. Ranked importance for each model is reported for each bar.

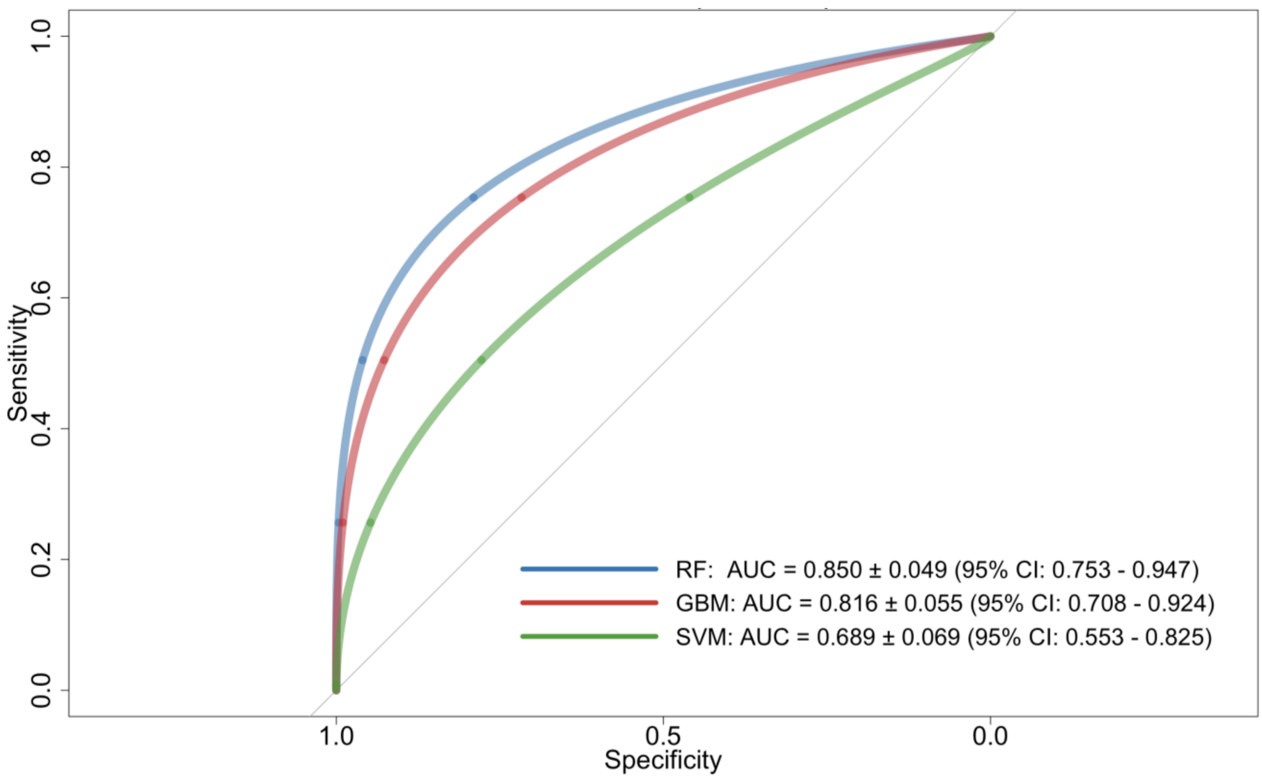


**Figure S3. Receiver operating characteristic (ROC) curves showing the classification performance of the three Machine Learning (ML) models evaluated in this study.** Model performance was quantified using the area under the ROC curve (AUC), a threshold-independent metric that measures the ability of a classifier to discriminate between classes. The Random Forest (RF) model achieved the highest performance (AUC = 0.85), followed by the Gradient Boosting Machine (GBM; AUC = 0.82), while the Support Vector Machine (SVM) showed lower discriminative performance (AUC = 0.69).

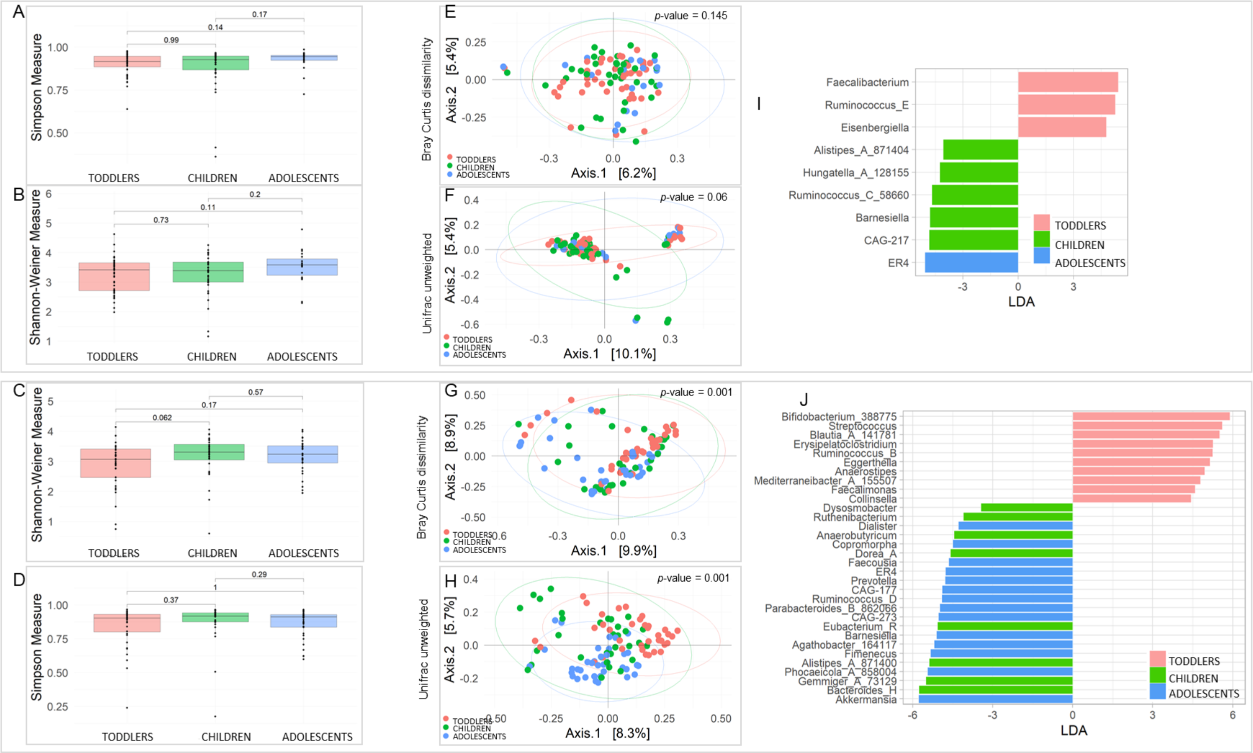

**Figure S4. Age-stratified gut microbiota in ASD patients and in CTRL.** Boxplots comparing Shannon diversity (**A and C**) and Simpson index diversity (**B and D**) in ASD and in CTRL age groups. Wilcoxon rank-sum p-values are reported. Bray–Curtis PCoA plot **(E and G)** and Unweighted UniFrac PCoA plot **(F and H)** of ASD and of CTRL age groups. PERMANOVA p values are shown. I and J: LEfSe analysis plot for ASD **(I)** and CTRL **(J)** age group comparisons.

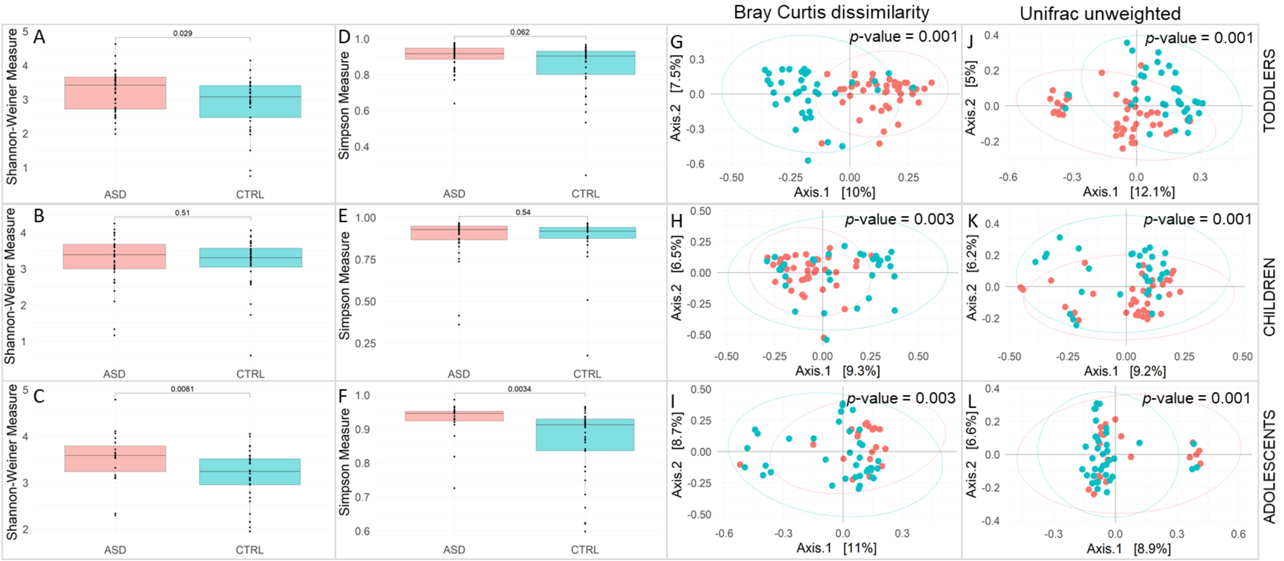


**Figure S5. Age-stratified gut microbiota in ASD patients compared to CTRL subjects.** Boxplots comparing Shannon diversity **(A-C)** and Simpson index diversity **(B-D)** in ASD versus CTRL in different age groups. Wilcoxon rank-sum p-values are reported. Bray–Curtis PCoA plot **(G-I)** and Unweighted UniFrac PCoA plot **(J-L)** of ASD versus CTRL in different age groups. PERMANOVA p values are shown.

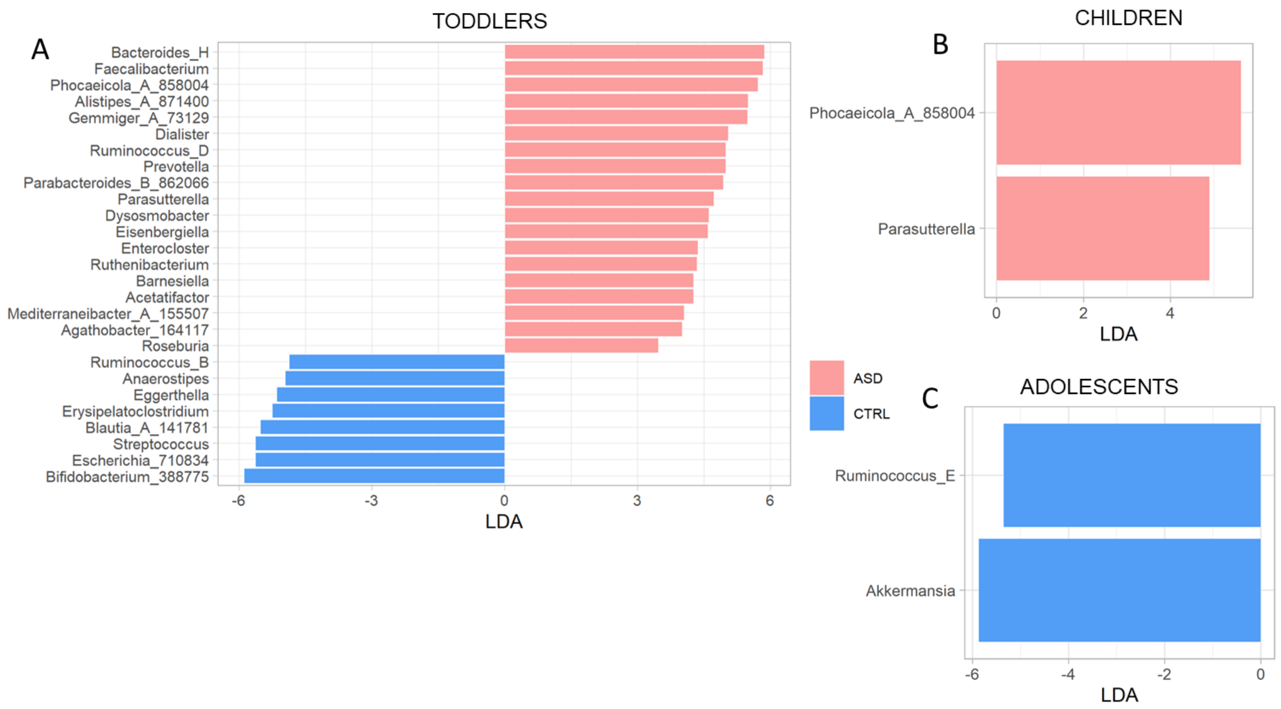


**Figure S6. Differential taxa composition between ASD and CTRL within each age group.** LEfSe analysis plot for ASD versus CTRL in toddlers group **(A)**; children group **(B)**; adolescents group **(C)**.

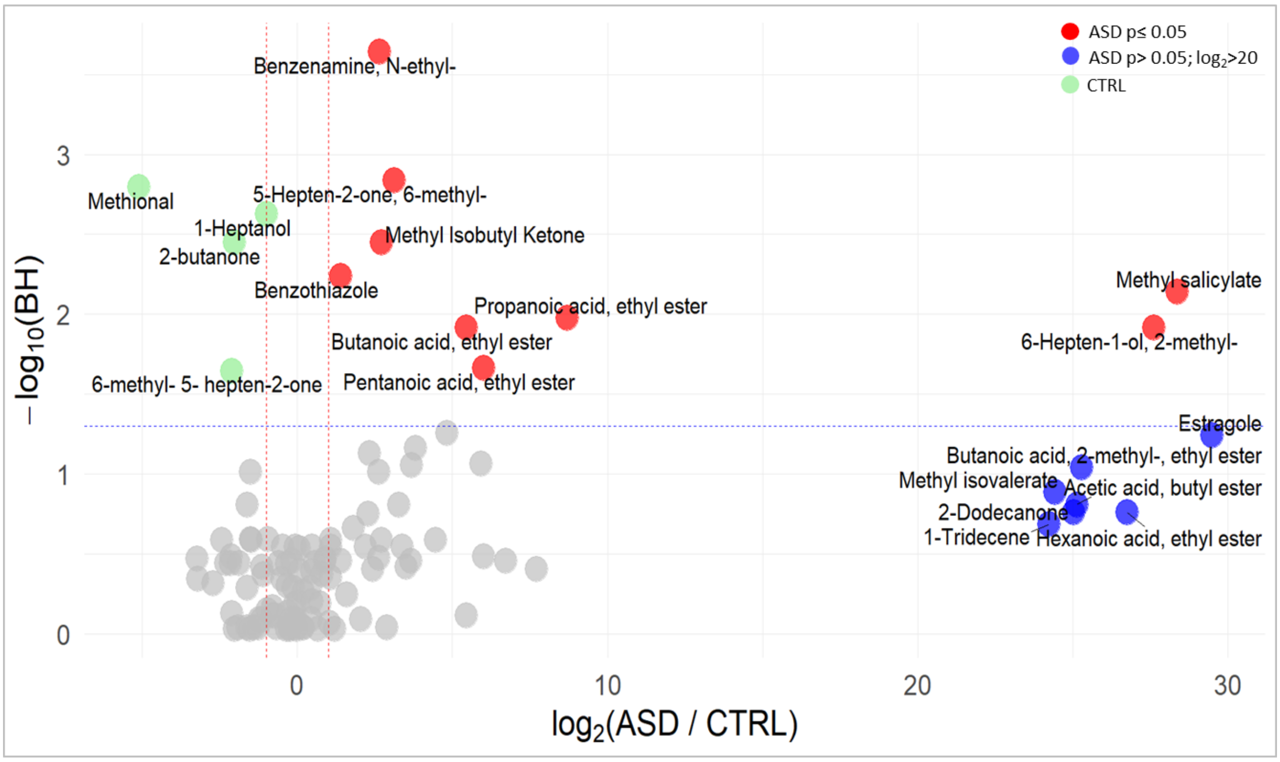


**Figure S7**. **Volcano plot of metabolites differentiating ASD and CTRL groups.** The volcano plot shows the distribution of all detected volatile organic compounds (VOCs) in stool samples, comparing ASD to CTRL subjects. Metabolites significantly dysregulated in ASD (adjusted $p < 0.05$, $|\log_2 FC| > 1$) are shown in red (higher in ASD) and green (higher in controls). Metabolites elevated in ASD but not reaching statistical significance are shown in blue.

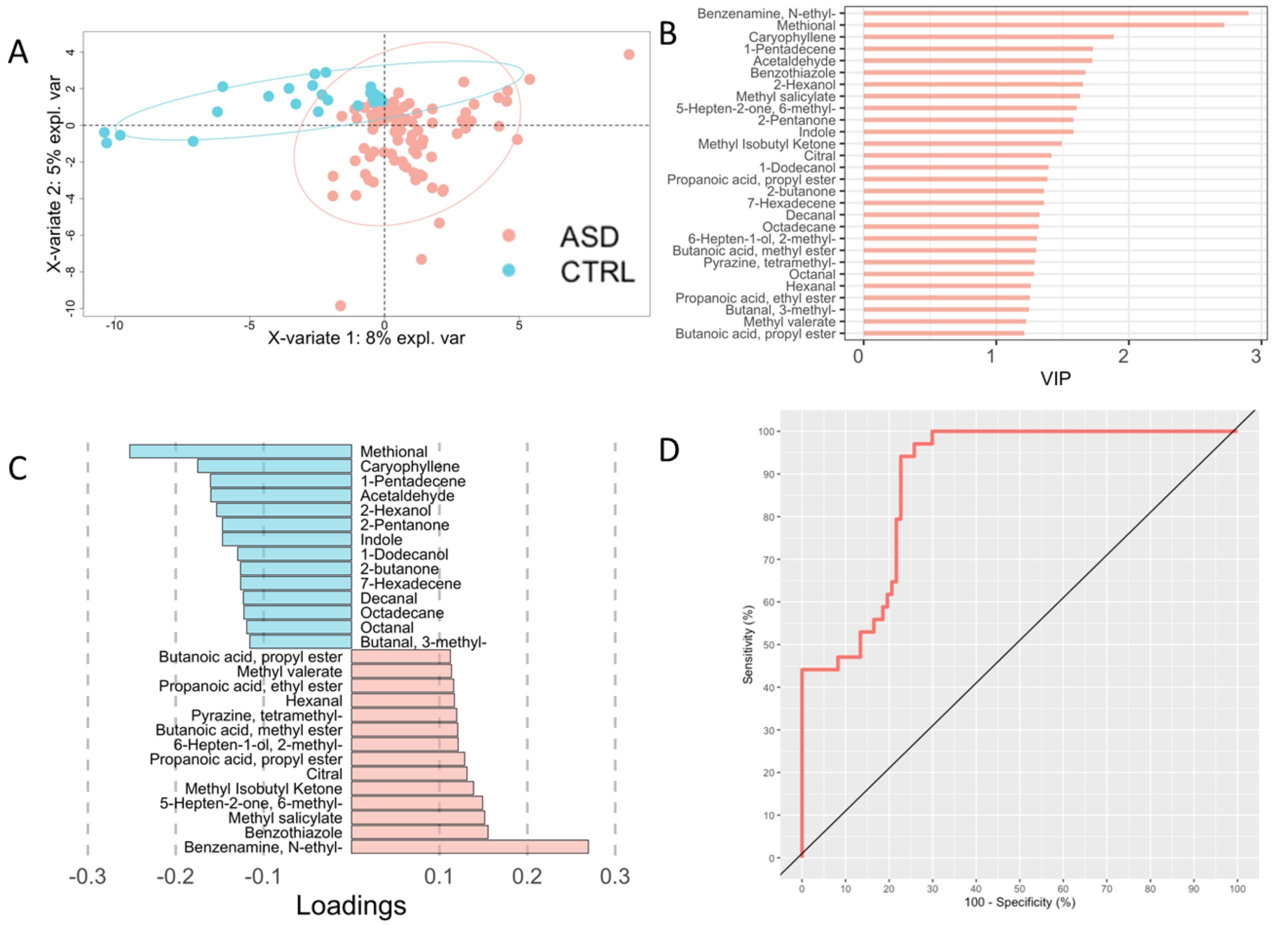


**Figure S8. PLS-DA model discriminating ASD from CTRL based on metabolomic profiles. (A)** PLS-DA demonstrates robust separation between ASD and CTRL groups **(A)**. VIP scores **(B)** and loading biplot **(C)** identify key discriminant metabolites contributing to classification accuracy. The model shows strong performance ($R^2 = 0.531$, $Q^2 = 0.279$, $p = 0.02$, RMSE = 0.305), with an AUROC of 0.886 **(D)**.

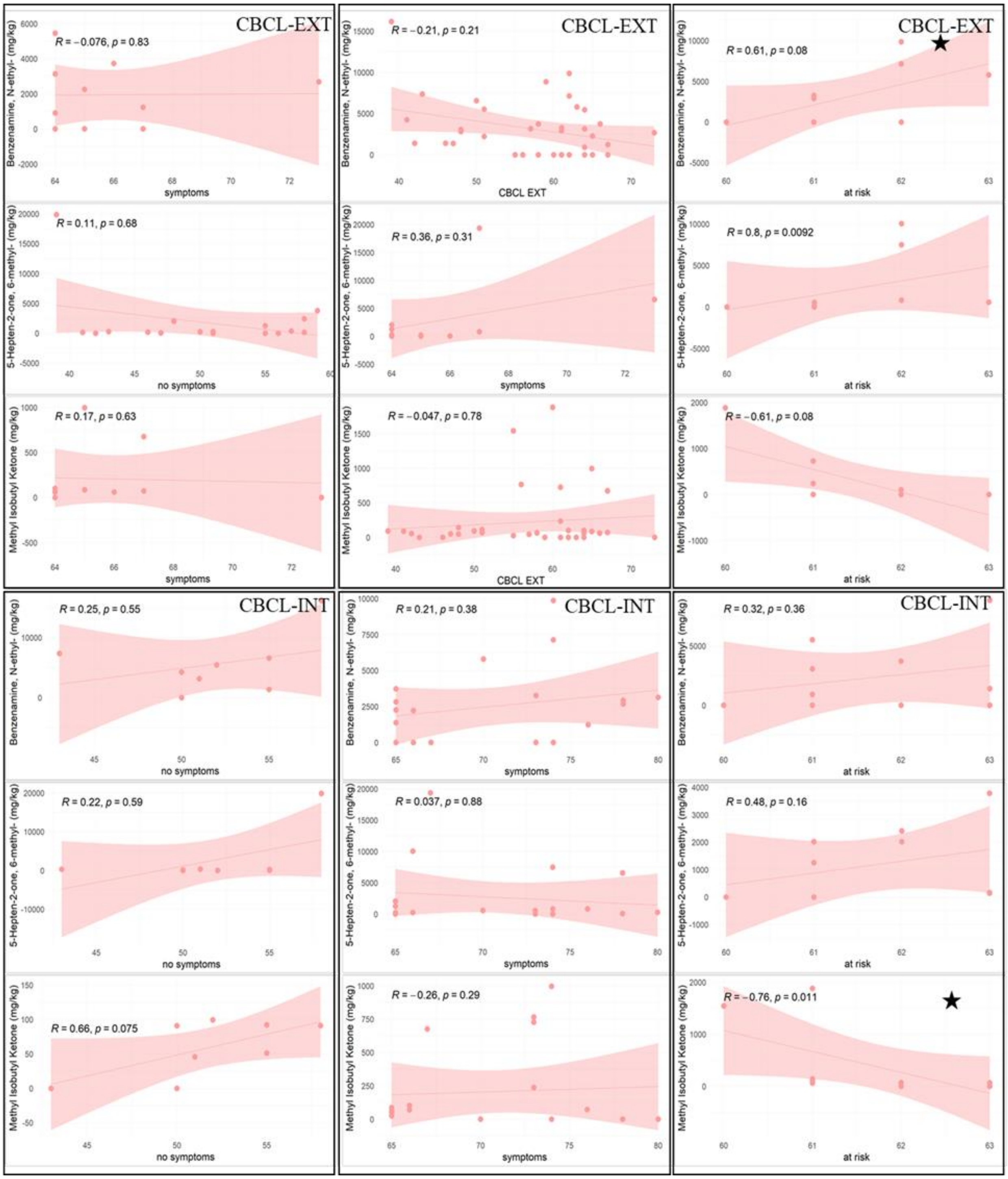


**Figure S9.** Linear regression analyses of machine learning–identified metabolites versus ASD Child Behaviour Checklist scale scores. Regressions with $p<0.05$ are highlighted by a star.

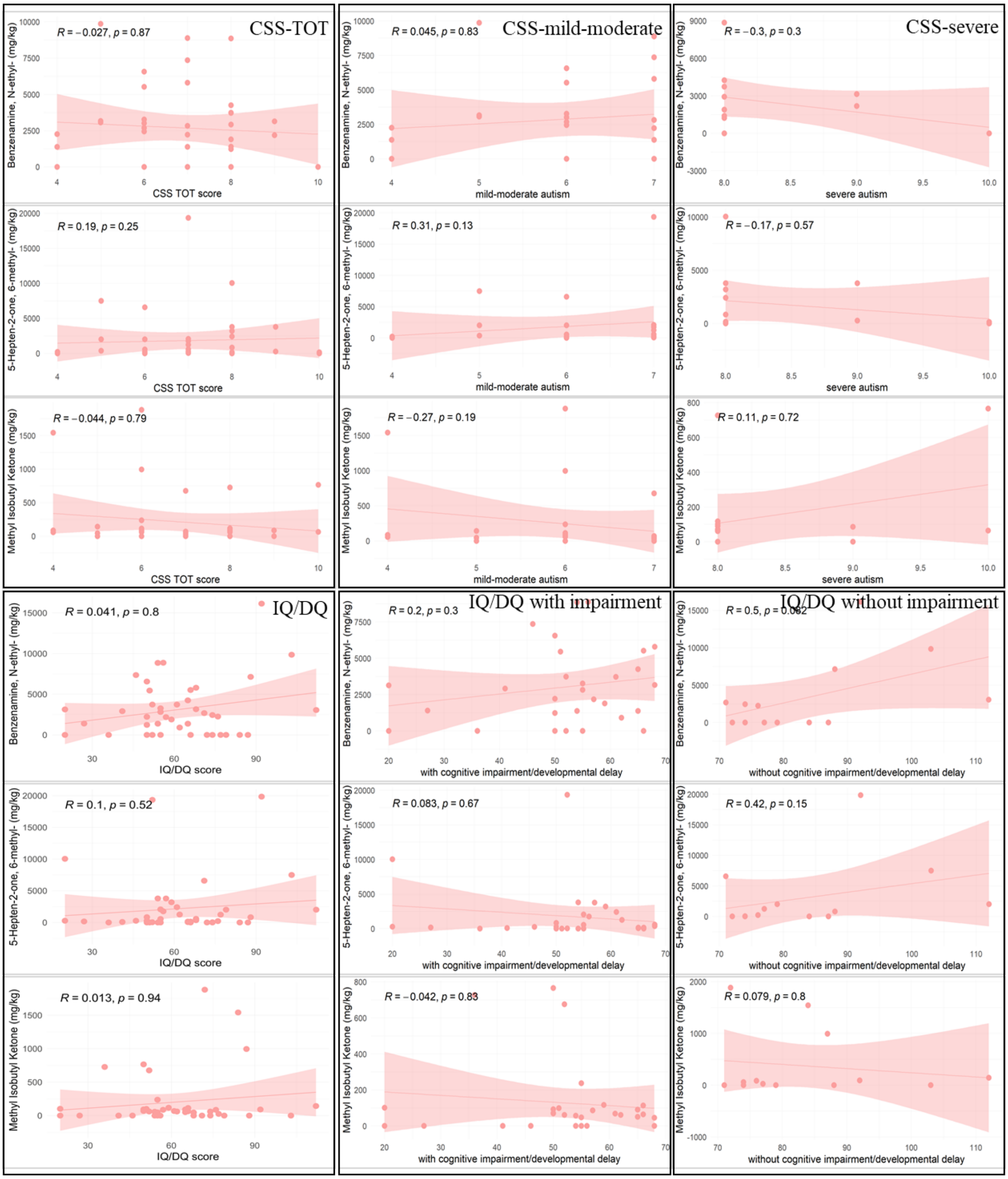


**Figure S10.** Linear regression analyses of machine learning–identified metabolites versus ASD Calibrated Severity Scores scale and intelligence quotient/developmental quotient score.

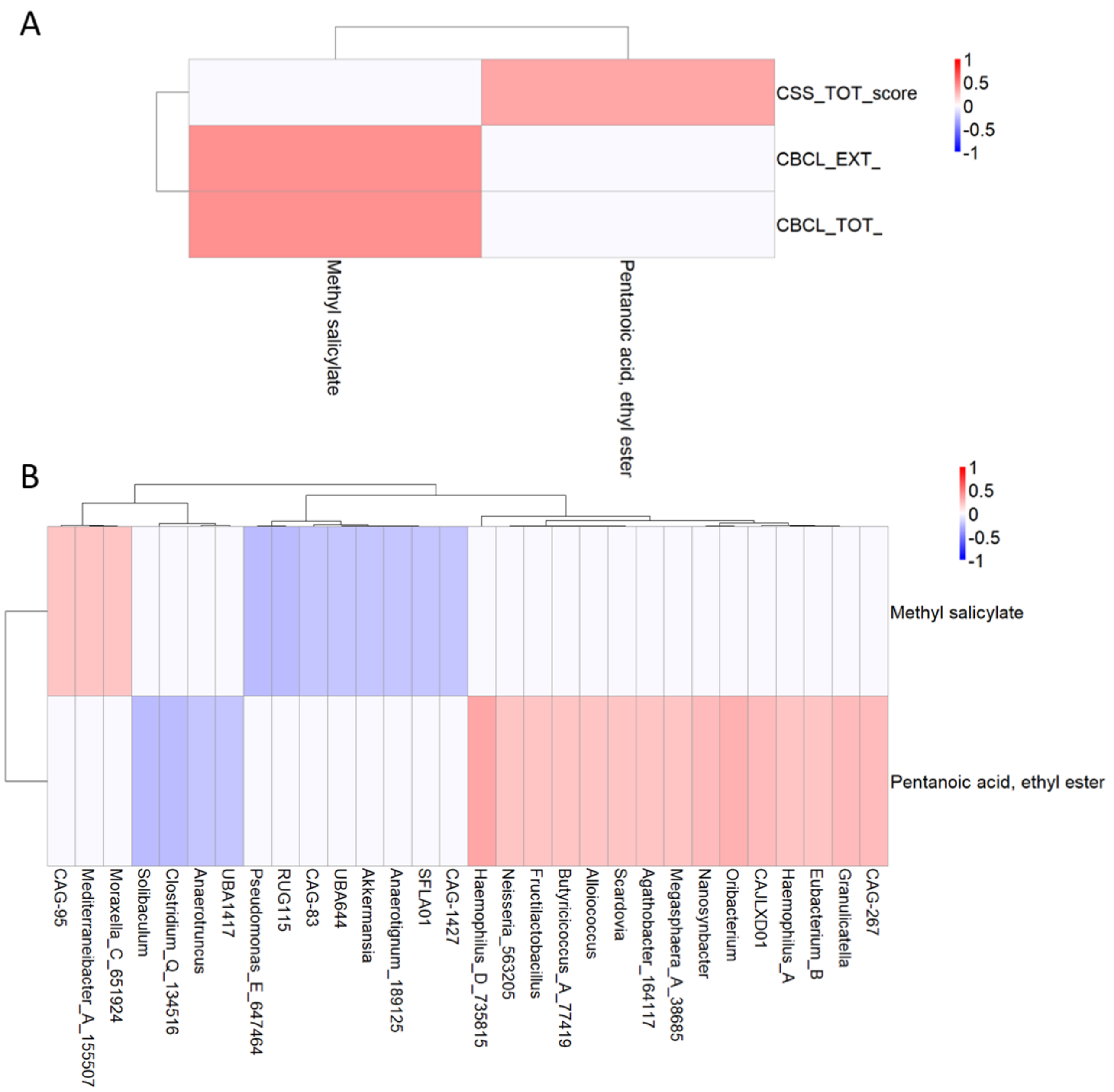


**Figure S11. Hierarchical clustered heatmaps of the association between VOCs and behavioural scales (A) and bacterial taxa (B).** Colours represent standardized association values (z-scores), ranging from blue (negative association) to red (positive association), as indicated by the colour scale on the right.

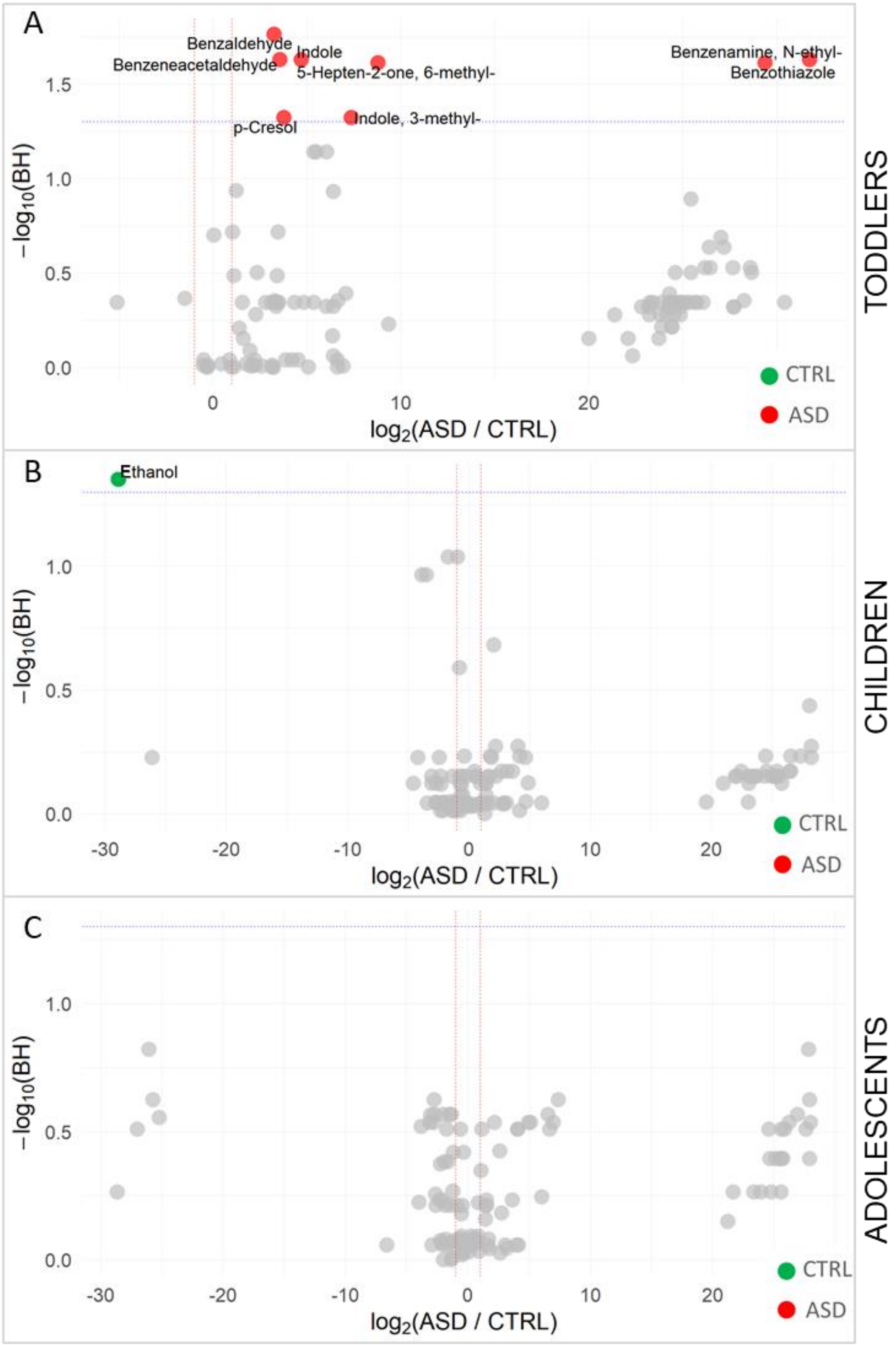


**Figure S12**. **Volcano plot of metabolites differentiating ASD vs CTRL within each age group.** The volcano plot shows the distribution of all detected volatile organic compounds (VOCs) in stool samples. Metabolites significantly dysregulated in ASD ($p_{adj} < 0.05$, $|log_2FC| > 1$) are highlighted.

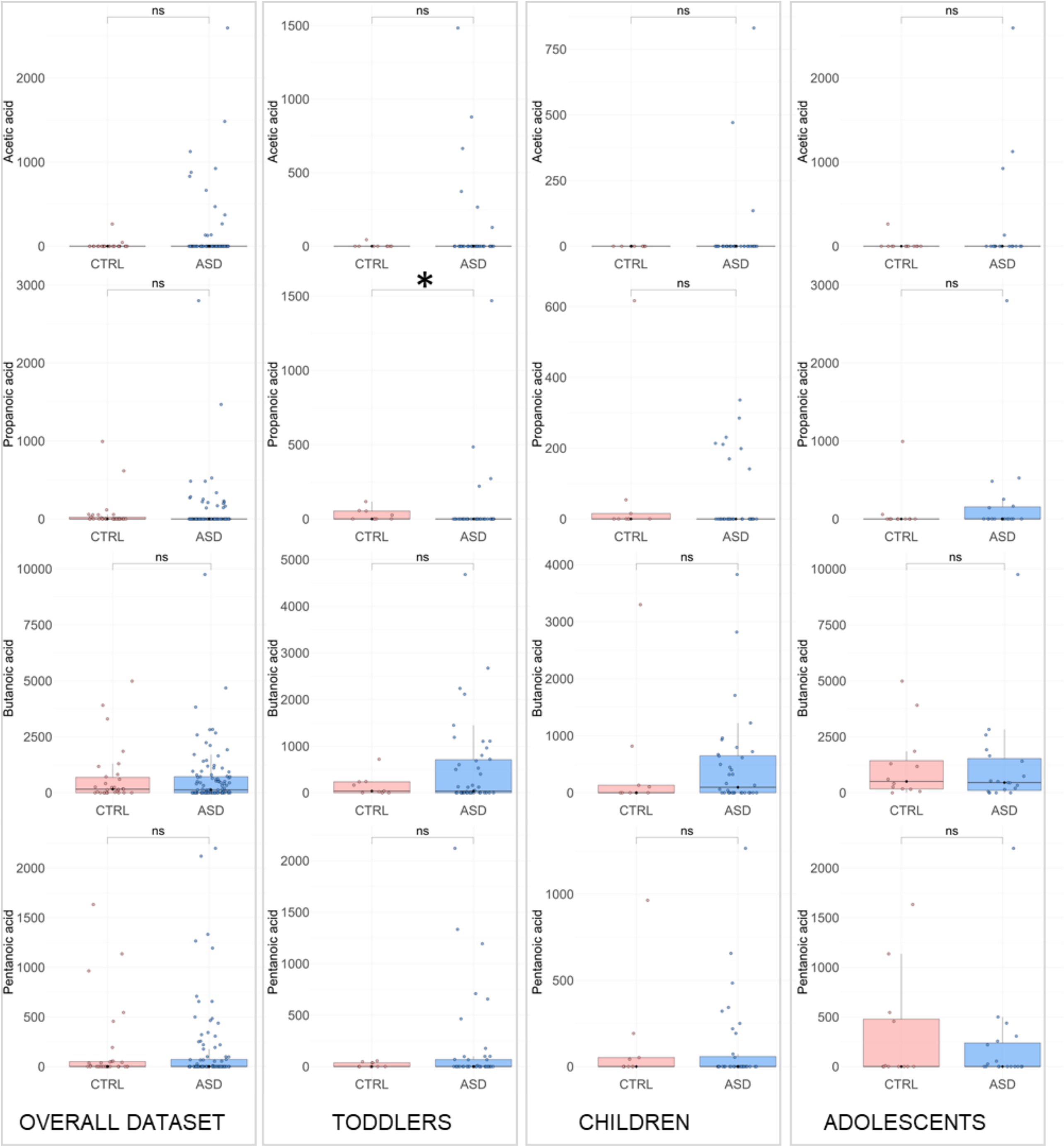


**Figure S13**. **Box plot of SCFA differentiating ASD vs CTRL and within each age group.** The box plot shows the distribution of acetic acid, propanoic acid, butanoic acid, pentanoic acid in stool samples. * comparisons statistically significant ($p_{adj} < 0.05$).

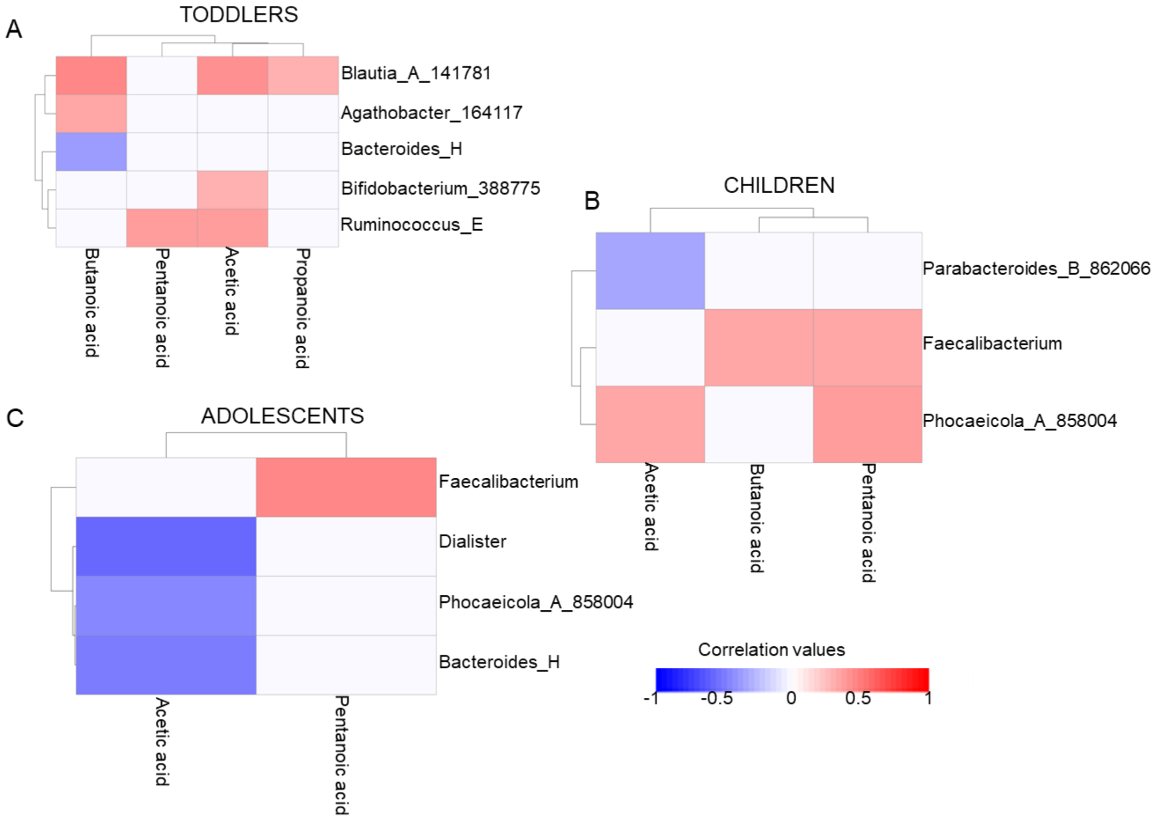


**Figure S14.** Correlations between bacterial taxa and short-chain fatty acids (SCFAs) in individuals with ASD, stratified by age.

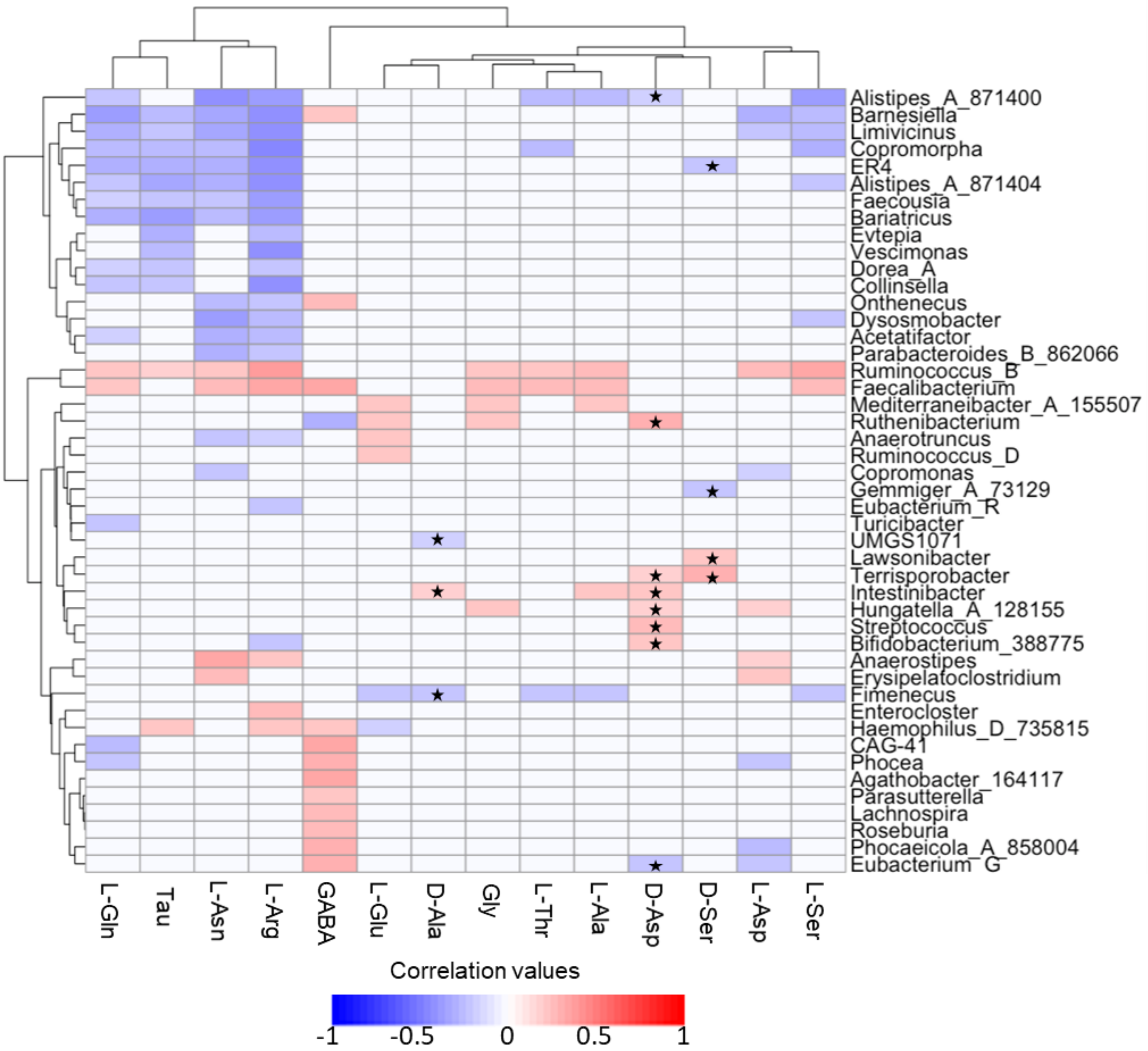


**Figure S15. Spearman correlation analysis of L- and D-AAs and bacterial taxa.** Positive values indicate direct correlations, while negative values indicate inverse correlations.

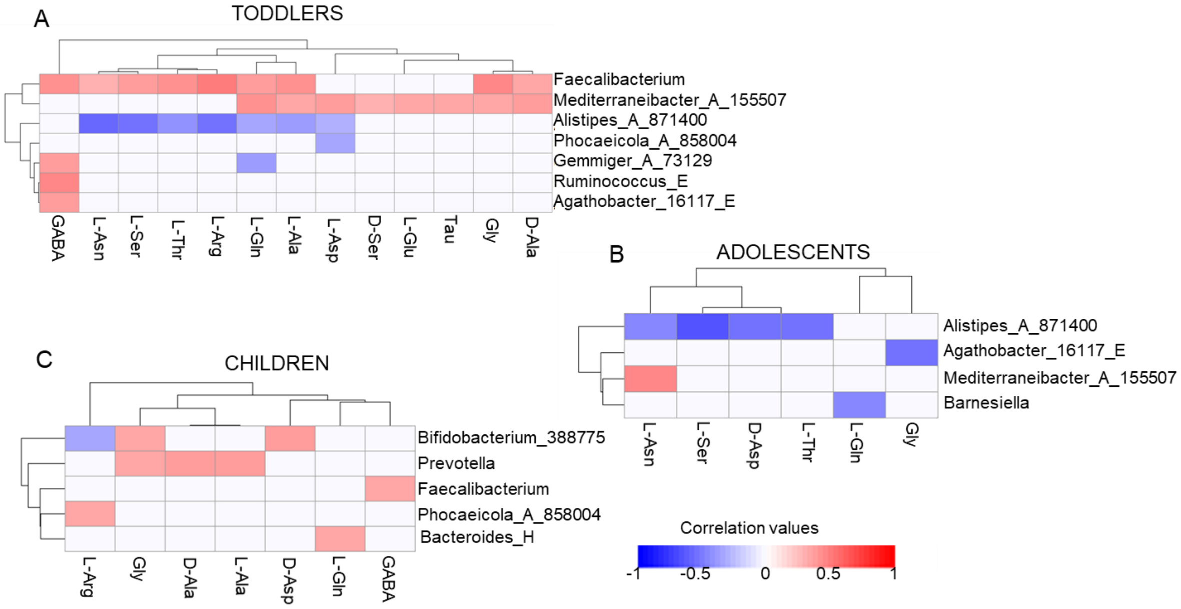


**Figure S16.** Age-stratified correlation analysis between faecal amino acids and bacterial taxa in ASD toddlers, ASD children, and ASD adolescents.

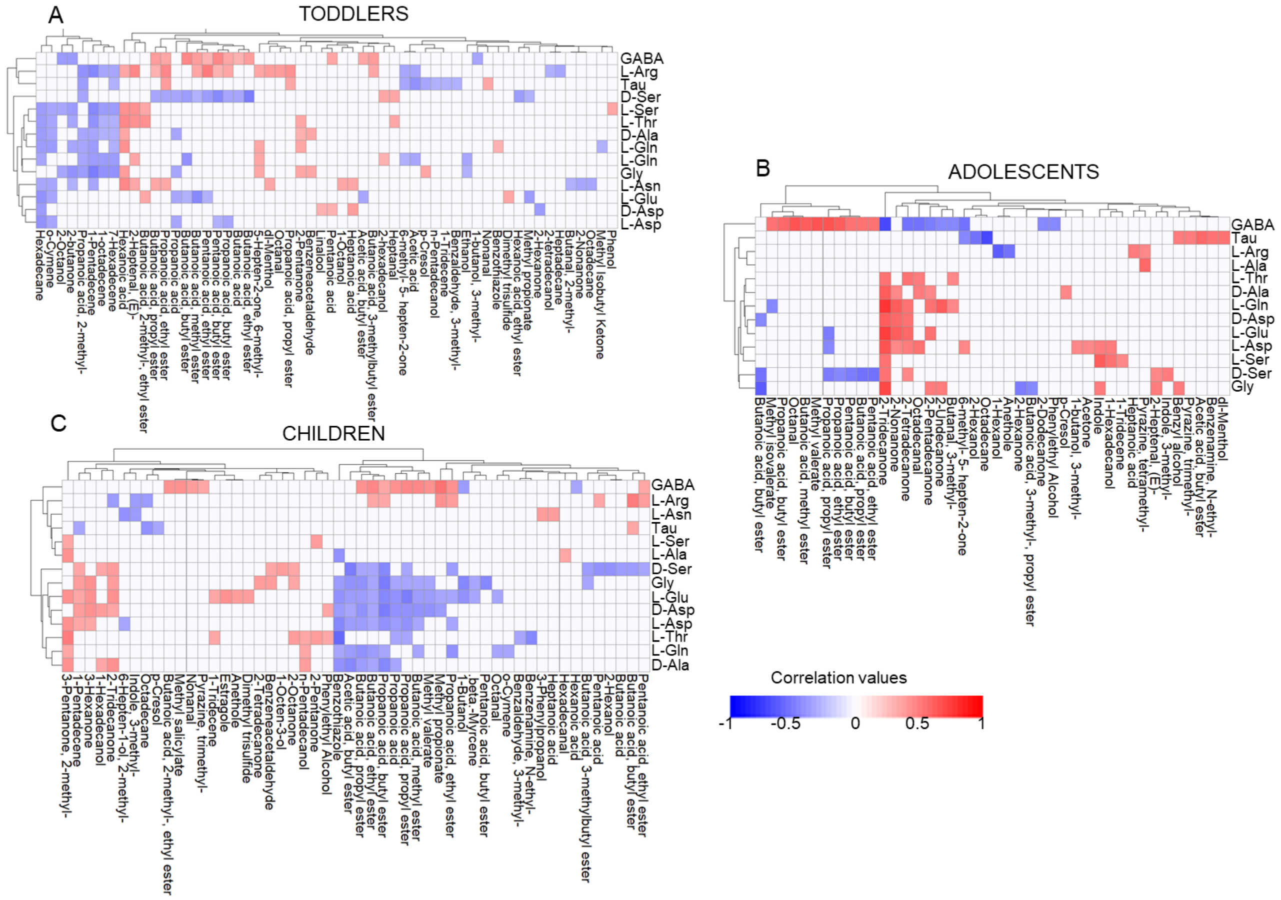


**Figure S17.** Age-stratified correlation analysis between faecal amino acids and metabolites in ASD toddlers, ASD children, and ASD adolescents.
